# Protein signatures associated with loneliness and social isolation: plasma proteome analyses in the English Longitudinal Study of Ageing, with causal evidence from Mendelian randomization

**DOI:** 10.1101/2024.07.25.24310989

**Authors:** Jessica Gong, Zohar Preminger, Andrew Steptoe, Daisy Fancourt

## Abstract

The understanding of biological pathways related to loneliness and social isolation remains incomplete. Cutting-edge population-based proteomics offers opportunities to uncover novel biological pathways linked to social deficits. This study employed a proteome-wide and data-driven approach to estimate the cross-sectional associations between objective measures of social connections (i.e., social isolation) and subjective measures (i.e., loneliness) with protein abundance, using the English Longitudinal Study of Ageing. Greater social isolation was associated with higher levels of 11 proteins (TNFRSF10A, MMP12, TRAIL-R2, SKR3, TNFRSF11A, VSIG2, PRSS8, FGFR2, KIM1, REN, and NEFL) after minimal adjustments; and three proteins were significantly associated after full adjustments (TNFRSF10A, TNFRSF11A, and HAOX1). Findings from two-sample Mendelian randomization indicated that a lower frequency of in-person social contact with friends or family causally increased levels of TNFRSF10A, TRAIL-R2, TNFRSF11A, and KIM1, and decreased the level of NEFL. The study also highlighted several enriched biological pathways, including necrosis and cell death regulation, dimerization of procaspase-8, and inhibition of caspase-8 pathways, which have previously not been linked to social deficits. These findings could help explain the relationship between social deficits and disease, confirming the importance of continuing to explore novel biological pathways associated with social deficits.

## INTRODUCTION

A large body of work now demonstrates the substantial impact that deficits in social connections have on health.[1, 2] Both social isolation (the objective presence or absence of other people in our lives and the frequency that which we engage with them) and loneliness (the perceived availability of others to fulfil our personal needs) are clear predictors of incidence of physical diseases (e.g. cardiovascular disease, dementia and diabetes),[3–6] psychiatric disorders (including depression, anxiety and schizophrenia),[7, 8] age-related decline,[9, 10] and mortality (both through suicide and other causes).[11–13]

Numerous different theories provide direct and indirect explanations for why these effects occur. In relation to **direct pathways**, according to the social neuroscience model, *homo sapiens* is an inherently social species, relying on others for species survival. If humans find themselves socially isolated and therefore vulnerable, they no longer feel safe, and experience social stress and emotional symptoms of isolation (i.e. loneliness), resulting in a cascade of bio-behavioral effects being activated, all to increase preparedness for potential assaults.[14] These effects include an increased vigilance for social threats, less settled sleep (to avoid predation), decreased impulse control, increases in depressive symptomatology (to signal the need for support and connection), and a host of physiological changes indicating a hypervigilant stress response system (e.g. elevated vascular activity, heightened hypothalamic-pituitary-adrenal (HPA) axis activity, and enhanced immune activation).[15] However, while these neural and behavioral responses may increase the likelihood of short-term survival, if isolation becomes repeated or chronic, they can carry long-term costs, including increasing the risk of chronic illness. Biologically, the mechanisms that specifically link chronic social isolation to disease include (i) an overactive HPA axis resulting in receptor cells developing glucocorticoid resistance, leading to a greater susceptibility to inflammation (itself associated with adverse health conditions), (ii) dysregulation of the autonomic nervous system’s ability to regular cardiovascular activity (including decreased heart rate variability), (iii) changes in immunological responses (including increased secretion of inflammatory cytokines, growth factors and antibodies) (iv) reduced repair and restorative processes (such as decreased natural killer cell activity and slower wound healing), and (v) changes in brain structure and functioning (including increased brain atrophy and reduced neurogenesis).[16–19]

In addition, social isolation and loneliness may influence health outcomes via **indirect pathways**. According to the social control hypothesis, social networks tend to discourage poor health behaviors and encourage good health behaviors, thereby supporting the health of the group as a whole by promoting the health of the individuals within it.[20] When individuals become socially isolated, they lose this beneficial influence and are more likely to engage in adverse health behaviors, including substance use (e.g. alcohol, cannabis, opiates, and tobacco), reduced physical activity, poor diet (leading to increased risk of malnutrition, eating disorders, and consumption of ultra-processed foods), reduced adherence to health guidelines (e.g. reduced medication adherence, use of preventative healthcare screening, visits to primary health care services, and following of basic hygiene procedures like hand-washing), and fewer recuperative behaviors (e.g. leisure activities).[1, 18]

However, while both these direct and indirect pathways provide some biological plausibility to the link between social connections and adverse health outcomes, they may not provide a complete picture. There is a risk that as certain biological pathways between social deficit and disease become better understood, they can dominate research, and attempts to identify novel pathways decline. In recent years, with the increasing availability of high-throughput measurements of molecular phenotypes such as protein abundance, there are novel opportunities to substantially expand our understanding of the biological implications of social deficits. Notably, several studies have focused on the impact of social isolation on protein expression in rodents, with findings identifying some novel biological pathways previously not connected to social deficits.[21–23] But to date, there are no studies focusing on humans.

Consequently, in this study, we focused specifically on identifying a protein signature for loneliness and social isolation using large-scale novel proteomics data from a representative cohort study. Literature on the relationship between social deficits and disease incidence suggest distinct risks from both loneliness and isolation,[3, 24–27] and previous research involving individual proteins relating to inflammatory pathways have corroborated the concept of distinct biological pathways for different types of social deficits.[28, 29] Therefore, we hypothesized that there would be distinct protein signatures for loneliness and social isolation. Our objectives were (i) to identify proteins associated with loneliness and social isolation, (ii) to apply two-sample Mendelian Randomisation (MR) to explore the potential causal effects of social deficits on the significantly associated proteins, and (iii) to undertake enrichment analysis of the identified proteins to identify which biological pathways are regulated by loneliness and/or isolation.

## RESULTS

### Study sample

At study wave 4 (2008-2009) in English Longitudinal Study of Ageing (ELSA), the mean score for loneliness was 4.3 [standard deviation (SD) = 1.6], while the mean score for social isolation was 2.7 [SD = 1.6] (Table 1), based on the pooled imputed datasets. The mean age of the study sample was 63.4 years [SD = 9.2]), 55% of the participants were female, and 97% were of White ethnicity.

**Table 1.** Descriptive statistics of participants in the sample.

|  | Raw dataset | Composite of 30 imputed datasets |
| --- | --- | --- |
| **Loneliness (scale 3-9)** |  |  |
| Mean (SD) | 4.2 (1.5) | 4.3 (1.6) |
| Missing | 384 (11.8%) | 0 (0.0%) |
| **Social Isolation (scale 0-7)** |  |  |
| Mean (SD) | 2.7 (1.6) | 2.7 (1.6) |
| Missing n (%) | 118 (3.6%) | 0 (0.0%) |
| **Age** |  |  |
| Mean (SD) | 63.4 (9.2) | 63.4 (9.2) |
| **Sex, n (%)** |  |  |
| Female | 1793 (55.0%) | 1793 (55.0%) |
| Male | 1469 (45.0%) | 1469 (45.0%) |
| **Ethnicity, n (%)** |  |  |
| White | 3163 (97.0%) | 3163 (97.0%) |
| Other ethnic groups | 99 (3.0%) | 99 (3.0%) |
| **Body Mass Index** |  |  |
| Mean (SD) | 28.2 (5.2) | 28.2 (5.3) |
| Missing n (%) | 81 (2.5%) | 0 (0.0%) |
| **Drinking (number of days with a drink over past 7 days)** |  |  |
| Mean (SD) | 2.2 (2.4) | 2.2 (2.4) |
| Missing n (%) | 22 (0.7%) | 0 (0.0%) |
| **Smoking Status, n (%)** |  |  |
| Never smoked | 1317 (40.4%) | 1327 (40.7%) |
| Formerly smoked | 1468 (45.0%) | 1478 (45.3%) |
| Currently smoking | 454 (13.9%) | 457 (14.0%) |
| Missing | 23 (0.7%) | 0 (0.0%) |
| **Education, n (%)** |  |  |
| Never went to school | 16 (0.5%) | 16 (0.5%) |
| Left school at 14 or under | 273 (8.4%) | 273 (8.4%) |
| Left school at 15 | 1026 (31.5%) | 1026 (31.5%) |
| Left school at 16 | 743 (22.8%) | 743 (22.8%) |
| Left school at 17 | 264 (8.1%) | 264 (8.1%) |
| Left school at 18 | 249 (7.6%) | 249 (7.6%) |
| Left school at 19 or over | 633 (19.4%) | 633 (19.4%) |
| Not yet finished school | 58 (1.8%) | 58 (1.8%) |
| **Non-Pension Wealth Quintiles, n (%)** |  |  |
| 1st | 718 (22.0%) | 737 (22.6%) |
| 2nd | 670 (20.5%) | 686 (21.0%) |
| 3rd | 611 (18.7%) | 624 (19.1%) |
| 4th | 630 (19.3%) | 642 (19.7%) |
| 5th | 560 (17.2%) | 573 (17.6%) |
| Missing | 73 (2.2%) | 0 (0.0%) |
| **Physical Activity, n (%)** |  |  |
| Less than monthly moderate and vigorous exercise | 424 (13.0%) | 424 (13.0%) |
| Monthly moderate and/or vigorous exercise | 196 (6.0%) | 196 (6.0%) |
| Weekly moderate or vigorous exercise | 1588 (48.7%) | 1588 (48.7%) |
| Weekly moderate and vigorous exercise | 1054 (32.3%) | 1054 (32.3%) |

### Associations between protein with loneliness and social isolation

First, the cross-sectional associations for loneliness and social isolation with the levels of 276 protein were estimated. By pooling the 30 imputed datasets, no protein was found to be significantly associated with loneliness based on the minimally-adjusted models (Figure 1). For social isolation, a total of 11 proteins were significantly associated after minimal adjustments, all indicating positive associations: **TNFRSF10A** (coefficient (β) [standard error (se)]: 0.061 [0.011]; False discovery rate (FDR) adjusted P (denoted as P_FDR_) = 9.89 × 10^-6^), **MMP12** (β [se]: 0.057 [0.011]; P_FDR_ = 4.10 × 10^-5^), **TRAIL-R2** (also known as **TNFRSF10B**) (β [se]: 0.056 [0.011]; P_FDR_ = 8.34 × 10^-5^), **SKR3** (also known as **ACVRL1**) (β [se]: 0.056 [0.011]; P_FDR_ = 0.0001), **TNFRSF11A** (β [se]: 0.056 [0.011]; P_FDR_ = 0.0001), **VSIG2** (β [se]: 0.049 [0.011]; P_FDR_ = 0.0026), **PRSS8** (β [se]: 0.045 [0.011]; P_FDR_ = 0.013), **FGFR2** (β [se]: 0.045 [0.011]; P_FDR_ = 0.021), **KIM1** (also known as **HAVCR1**) (β [se]: 0.043 [0.011]; P_FDR_ = 0.026), **REN** (β [se]: 0.041 [0.011]; P_FDR_ = 0.045), and **NEFL** (β [se]: 0.035 [0.009]; P_FDR_ = 0.047) (Figure 2). Based on the fully-adjusted models, there were no proteins significantly associated with loneliness (Supplementary Fig. 1). For social isolation, **TNFRSF10A** (β [se]: 0.049 [0.011]; P_FDR_ = 0.003), **HAOX1** (β [se]: 0.047 [0.012]; P_FDR_ = 0.014), and **TNFRSF11A** (β [se]: 0.044 [0.011]; P_FDR_ = 0.026) were found to be significantly associated in the fully-adjusted model (Supplementary Fig. 2), indicating higher protein levels associating with greater level of social isolation.

**Figure 1.**
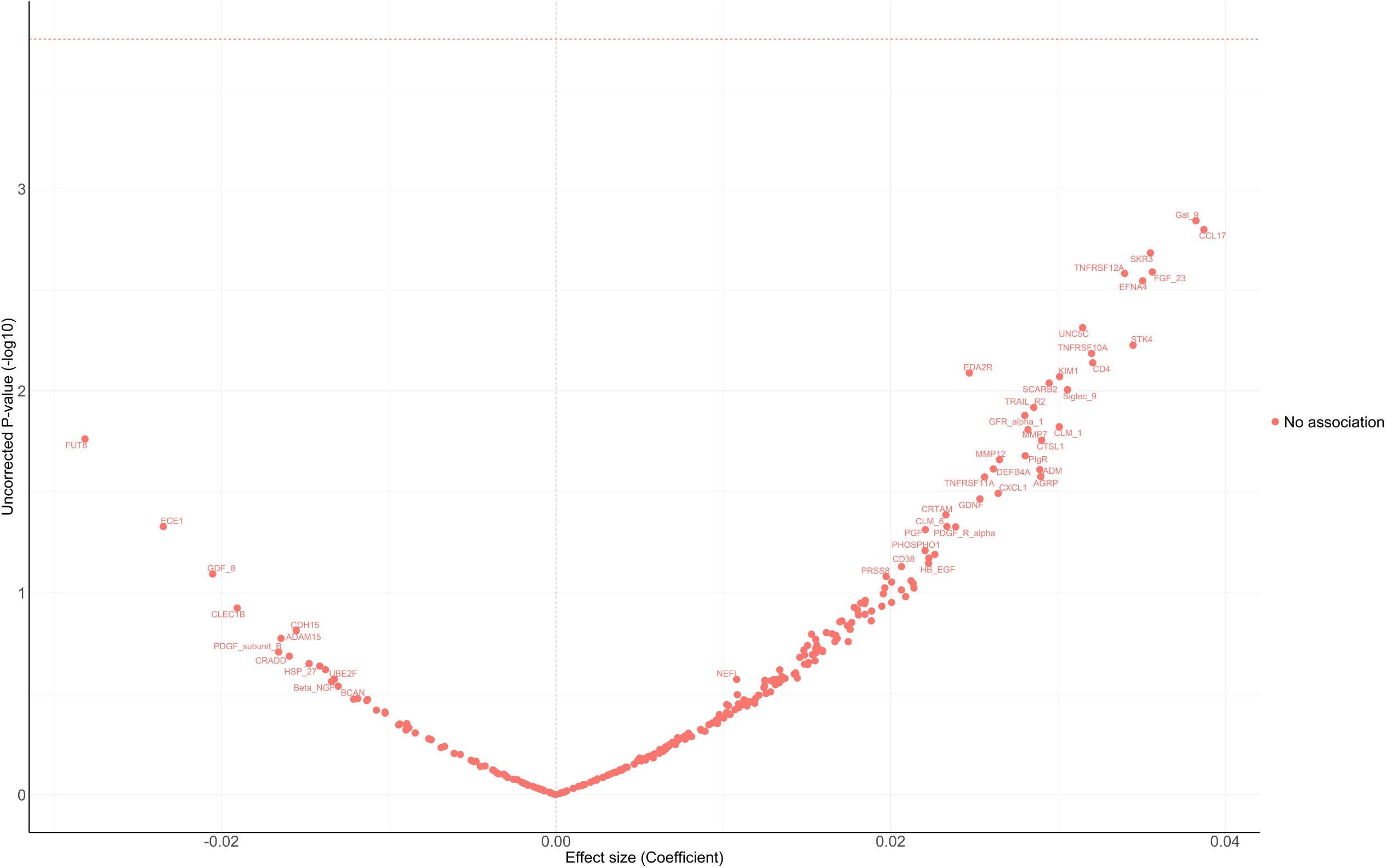
Volcano plot showing minimally adjusted coefficient (x axis) and uncorrected two-sided P values (y axis) for the association between protein concentration with loneliness using imputed data. Coefficient from linear regression models adjusted for age, sex, education, ethnicity, education, wealth quintile. Proteins above the horizontal dotted red line were significantly associated with loneliness after false discovery rate (FDR)-correction with p-value < 0.05.

**Figure 2.**
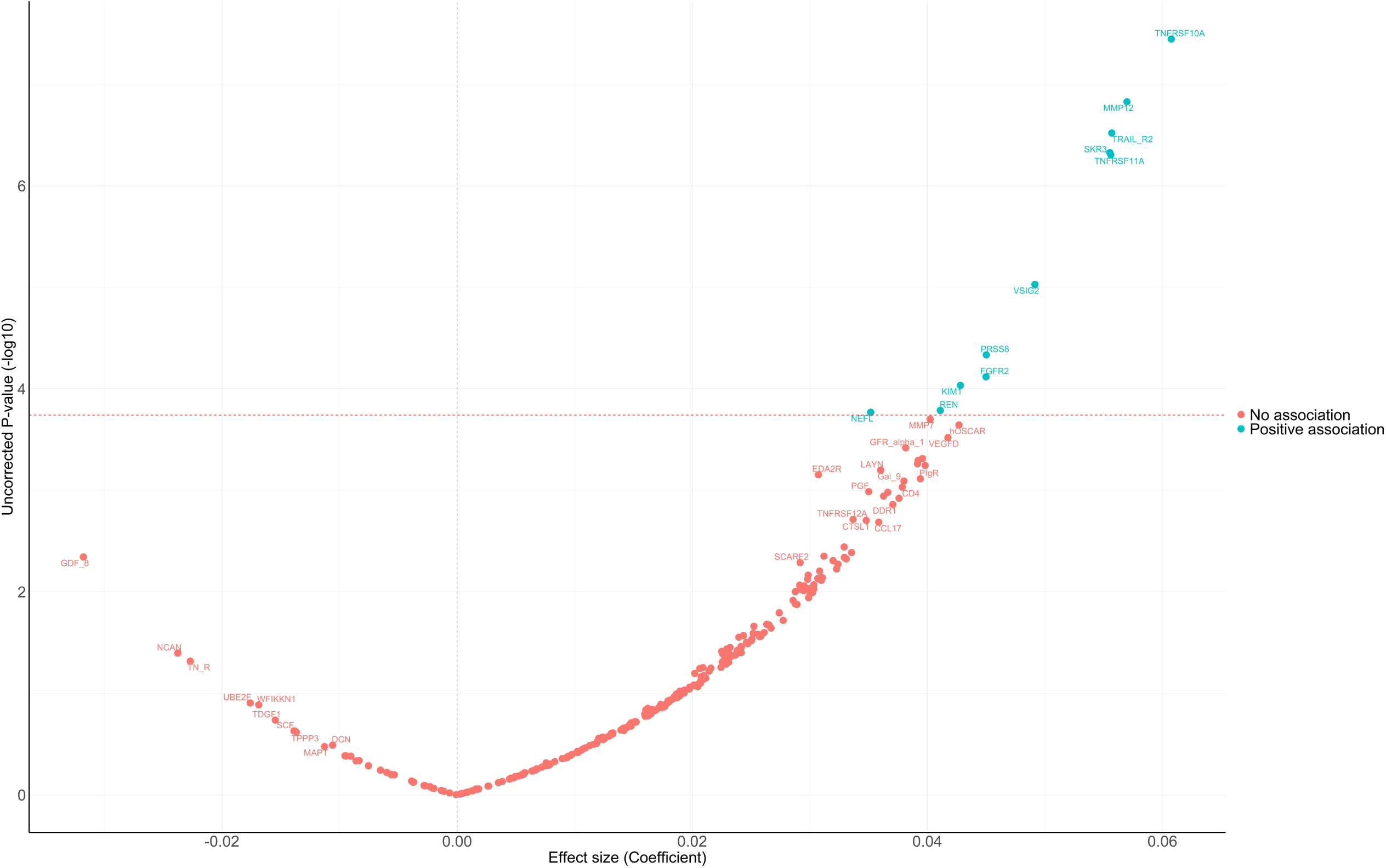
Volcano plot showing minimally adjusted coefficient (x axis) and uncorrected two-sided P values (y axis) for the association between protein concentration with social isolation using imputed data. Coefficient from Linear Regression models adjusted for age, sex, education, ethnicity, education, wealth quintile. Proteins above the horizontal dotted red line were significantly associated with social isolation after false discovery rate (FDR)-correction with p-value < 0.05.

### Two-sample MR

Next, two-sample MR was conducted using summary statistics from genome-wide association study (GWAS) to assess the potential causal relationship between social isolation and the protein concentrations identified as significant based on the linear regressions. Summary statistics for genetic variants associated with circulating protein levels, specifically protein quantitative trait loci (pQTL), which are also associated with dimensions of social isolation in the GWAS from the UK Biobank, were used to infer causality.

Based on results from two-sample MR, lower frequency of friend/family visits was causally linked to an increased level of **TNRFSF10A** (β [se]: 0.245 [0.117]; P = 0.037, based on IVW; β [se]: 0.324 [0.143]; P = 0.023, based on weighted median; β [se]: 0.249 [0.103]; P = 0.016, based on Maximum likelihood) (Figure 3; Supplementary Table 1); and increased level of **TRAIL-R2** (also known as **TNRFSF10B**) (β [se]: 0.264 [0.102]; P = 0.010, based on IVW; β [se]: 0.274 [0.104]; P = 0.008, based on Maximum likelihood); increased level of **TNRFSF11A** (β [se]: 0.210 [0.103]; P = 0.042, based on Maximum likelihood); increased level of **KIM1** (also known as **HAVCR1**) (β [se]: 0.195 [0.095]; P = 0.040, based on Maximum likelihood); and decreased level of **NEFL** (β [se]: −0.257 [0.105]; P = 0.014, based on IVW; β [se]: −0.248 [0.101]; P = 0.014, based on Maximum likelihood). MRs of living alone or the multi-trait loneliness-isolation GWAS did not show any significant findings.

**Figure 3.**
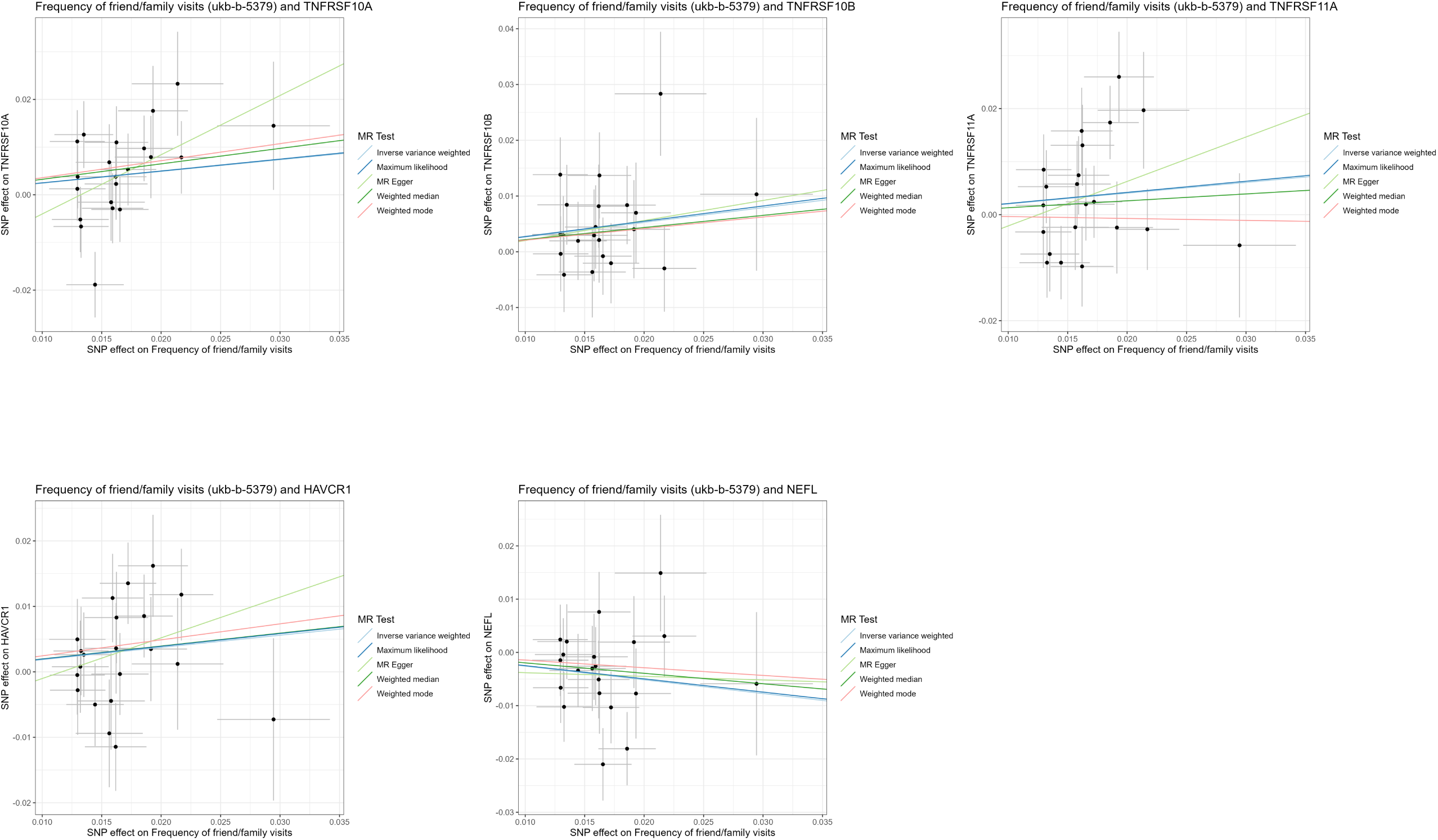
Two-sample Mendelian randomisation scatter plots for frequency of friend/family visits and TNFRSF10A, TRAIL-R2 (TNFRSF10B), TNFRSF11A, KIM1 (HAVCR1), NEFL. Analyses were conducted using the inverse variant weighted, maximum likelihood, MR-Egger, Weighted median, Weighted mode methods. The slope of each line corresponding to the estimated MR effect per method.

### Enrichment analysis

The enrichment analysis of the identified proteins unveiled a spectrum of pathways and expressions significantly enriched, by searching relevant public bioinformatics databases using EnrichR [30] (Figure 4; Supplementary Table 2). These pathways include RIPK1-mediated regulated necrosis, TP53 regulation of death receptors and ligands transcription, and TRAIL signalling. Additionally, pathways regulating necrosis and cell death, such as death receptor activity and caspase activation via death receptors in the presence of ligands, and via extrinsic apoptotic signalling pathways, were prominently featured. Moreover, inhibition of caspase-8 activity, and dimerization of procaspase-8 pathways was identified. These findings collectively indicate the identified proteins have pivotal role in regulating processes associated with cell death, death receptor signalling cascades, and caspase activation.

**Figure 4.**
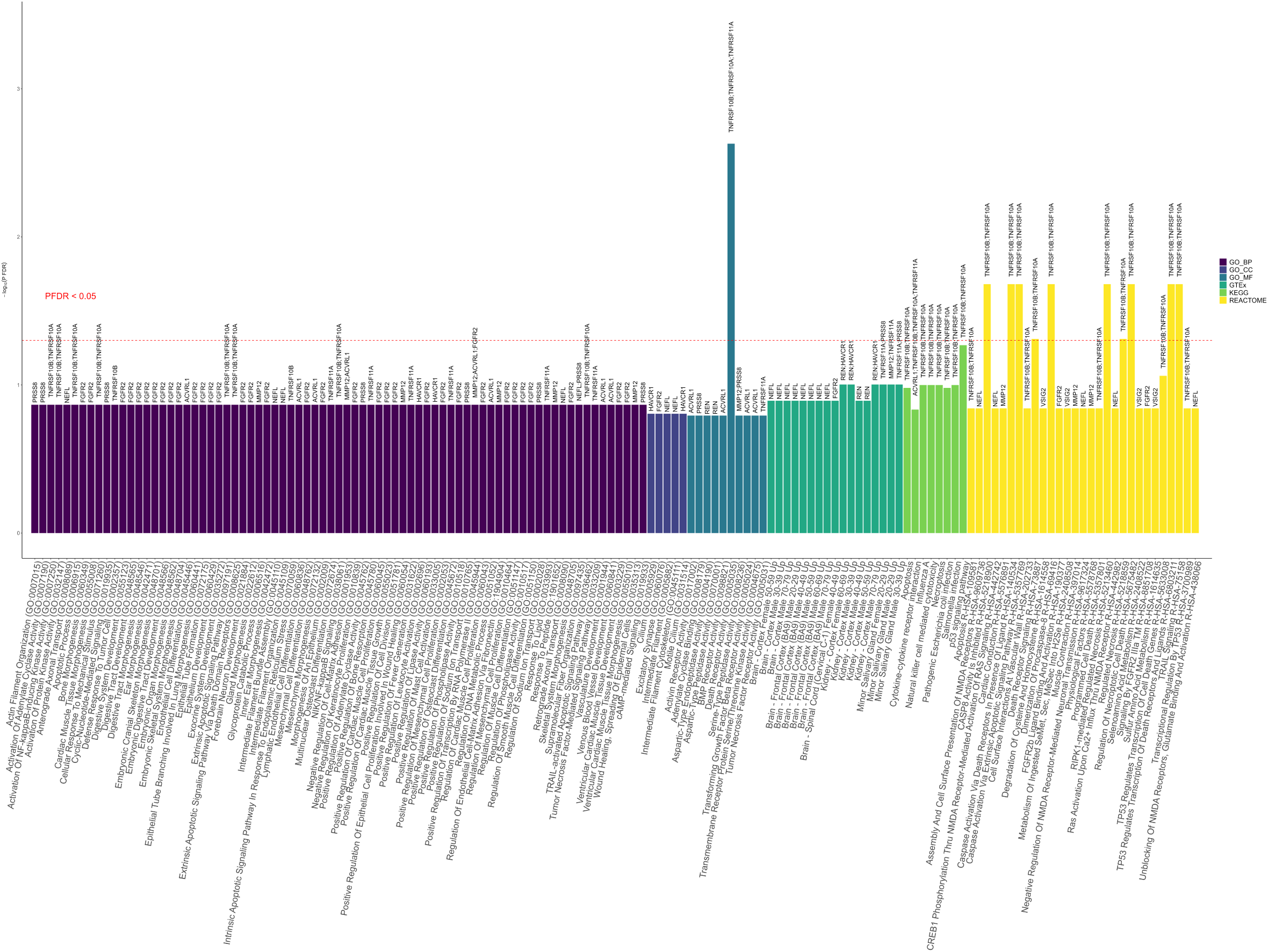
Enrichment analysis of the identified proteins. Enrichment for Gene Ontology (GO) 2023, Genotype-Tissue Expression (GTEx) 2023, Kyoto Encyclopaedia of Genes and Genomes (KEGG) 2021, and Reactome pathways 2022. Significant proteins after false discovery rate (FDR) correction derived from linear regression models in the minimally- and fully-adjusted models were fed into Enrichr (https://maayanlab.cloud/enrichr/) for enrichment analysis. The full list of proteins from ELSA was used as the background gene set. Terms displayed on the bar plot were filtered by a two-sided p-value < 0.05. Terms above the horizontal dotted red line were enriched after FDR-correction with p-value < 0.05, and the text were highlighted in red.

## DISCUSSION

Overall, the current study utilized a data-driven approach to reveal a protein signature for social deficits. As hypothesized, we saw distinct findings for objective measures of social connections (i.e. social isolation) and subjective measures (i.e. loneliness). Specifically, greater social isolation was related to increased levels of 11 proteins cross-sectionally (**TNFRSF10A, MMP12**, **TRAIL-R2** (also known as **TNFRSF10B**), **SKR3** (also known as **ACVRL1**), **TNFRSF11A**, **VSIG2**, **PRSS8**, **FGFR2**, **KIM1** (also known as **HAVCR1**), **REN**, and **NEFL**), after minimal adjustments; and three proteins were significantly associated after full adjustments (**TNFRSF10A**, **TNFRSF11A**, and **HAOX1**). Findings from two-sample MR indicated that a lower frequency of in-person social contact with friends or family causally increased levels of **TNRFSF10A, TRAIL-R2** (also known as **TNFRSF10B**), **TNRFSF11A,** and **KIM1** (also known as **HAVCR1**), and decreased level of **NEFL**. The study also highlighted several enriched biological pathways, including necrosis and cell death regulation, dimerization of procaspase-8 and inhibition of caspase-8 pathways, which have previously not been linked to social deficits and could help to explain their relationship to disease, confirming the importance of continuing to explore novel biological pathways associated with social deficits.

This is, to our knowledge, the first study looking at the relationship between phenotypic loneliness and social isolation and human protein expression. A previous study mapped protein expression to GWAS that had been undertaken to identify single nucleotide polymorphisms (SNPs) associated with composite indices of social deficits (loneliness, living alone, low social contact and a lack of confidants).[31] There was no overlap between the proteins identified there and in our study. However, that study explored the alternative end of the molecular cascade, i.e. how genetic risk for loneliness or isolation is related to protein expression, rather than how phenotypic experiences of loneliness or isolation could moderate protein expression. What the combination of our findings with those of this previous study suggest is that the molecular influence of phenotypic experiences of loneliness/isolation may not be restricted to proteins transcribed and translated from genes specific to loneliness/isolation. Indeed, even looking across other genetics studies, none of the proteins that related to social isolation come from genes that have previously been related to social deficits.[32]

Instead, we identified novel proteins that may be causally influenced by social isolation, and in doing so, we identify a number of new biological pathways that have previously not been associated with social deficits. TNFRSF10A, TNFRSF10B (also known TRAIL-R2) and TNFRSF11A (also known as TRANCE receptor or RANK) are upregulated in the blood in various cancers including myeloma, several leukaemia and lung cancer, which could provide some explanation for the relationship between isolation and cancer mortality.[25] TNFRSF10A and TNFRSF10B also initiate the cascade of caspases that mediates apoptosis and promotes the activation of NF-κB.[33] According to the Conserved Transcriptional Response to Adversity theory (CTRA), when socially isolated, humans are evolutionarily primed to express a higher concentration of genes relating to increased inflammation and decreased antiviral activity.[34] The social signal transduction process involved in this gene expression is thought to involve peripheral neural signalling via neurotransmitters such as dopamine and norepinephrine that then leads to cellular signal transduction and the activation of multiple transcription factors including CREB and NF-κB. So, our finding of greater TNFRSF10A and TNFRSF10B abundance in response to social isolation aligns with previous theoretical and empirical work.

Amongst other proteins that we identified as associated with social isolation, KIM1 (also known as HAVCR1) is a member of the TIM (T-cell immunoglobulin and mucin) gene family, which plays an important role in host-virus interactions, and acts as a receptor for viruses including Ebola, Marburg, Dengue, Zika and possible SARS-CoV-2.[35] This could help to explain studies demonstrating increased viral susceptibility amongst those who are socially isolated.[36] However, it is perhaps surprising that NEFL levels were lower amongst those with social isolation. NEFL is a biomarker of axonal damage and is widely implicated in neurological disorders. We also identified diverse enriched biological pathways related to cell death, death receptor signalling cascades, and caspase activation (intrinsic both to apoptosis and inflammation responses). This is noteworthy not just in terms of alignment with the literature on immunological response to social isolation, but also given oxi-inflamm-aging theories suggesting the critical role of chronic oxidative and inflammatory stress in biological ageing specifically.[37] However, caution is important when attempting to interpret the clinical relevance of these proteins. Given the highly complex interplay between molecular biomarkers in the pathogenesis of disease, further research is required to replicate the findings here in other datasets.

While we identified these biological responses for social isolation, it is notable that no proteins in our analyses were associated with loneliness in fully-adjusted models. In considering why we saw results for isolation but not loneliness, there are two explanations. Taking both a social neuroscience perspective and considering the social control hypothesis, it is the act of being physically isolated that is the risk to survival and activates direct and indirect pathways. However, it is also important to remember that this study focused on protein panels that were developed to be particularly relevant to neurological and cardiovascular conditions. Loneliness may be related to protein abundance at other parts of the human proteome. As such, our results do not necessarily imply that social isolation has greater effects on protein abundance than loneliness.

It is also of note that although we did identify a signature involving several different proteins associated with social isolation, MR analyses suggested the finding was not necessarily causal across all MR analyses. Some of the proteins identified in this study could be correlates of other molecular biomarker changes that occur due to social isolation or even of the disease states that are increased as a result of social deficits, rather than direct causal products of isolation themselves. Further, although we used large GWAS data on social isolation, none of the measures of isolation exactly replicated the social isolation phenotype we used, so there may be additional aspects of our phenotype not captured in the GWAS. Frequency of social contact (the MR that produced the findings discussed above) was the closest in measuring objective aspects of structural social connections. The “living alone” GWAS was limited in only measuring the more limited domain of domestic isolation, while the multi-trait GWAS included combinations of structural, functional and quality aspects of social connections that are much broader than the structural construct we tested phenotypically. Our GWAS summary data were also taken from UK Biobank, which contains a younger age range than ELSA, and age has been shown to affect the prediction accuracy of polygenic score (PGS).[38] As such, it is possible that alternative GWAS could have yielded different results. There are also broader acknowledged challenges around using MR for social and behavioral traits.[39] While a person’s genome can undoubtedly influence factors that have a relevance to social traits, from physical traits to personality characteristics, health factors and behaviors that could causally influence loneliness/isolation, the degree of variance that can be explained with a PGS is always limited by the true heritability a trait, which is likely smaller for complex social traits like loneliness/isolation that have large environmental components.[40] If the alleles in the PGS for loneliness/isolation are not directly causally related to loneliness/isolation but in fact based on indirect pathways (e.g. assortative mating of sociable parents) or the involvement of many pleiotropic genetic variants also strongly involved in downstream health conditions related to loneliness,[18] polygenic loneliness/isolation scores may in fact not be a valid tool to enhance our understanding of causality. As a result, we encourage that the MR analyses reported are interpreted cautiously.

This study has many strengths, including its use of a large, representative cohort of older adults of white European ancestry, rich validated measures of both social isolation and loneliness, and use of MR and enrichment analyses to explore issues of causality and biological plausibility with respect to disease risk. However, the study has several limitations. Because we focused on older adults, we do not know whether the biological responses identified in this study are stable across the life-course. We only had data on protein abundance for one timepoint, so we focused on cross-sectional relationships between social deficits, using MR to explore the direction of the association. But as the incorporation of high-throughput measurements of molecular phenotypes becomes more common within cohort studies, the potential inclusion of further waves of proteomics data could support the analysis of changes in protein abundance over time in relation to changing social deficits. Consequently, many pathways related to loneliness and social isolation might not have been detected. Broader proteome-wide studies are encouraged to replicate and extend our findings. Additionally, this study focused on the chronic effects of loneliness and social isolation on the proteome. But given animal research suggesting that isolation also induces acute fluctuations in protein expression, leading to different molecular signatures for each stage of response,[21] future studies are encouraged that explore the dynamic profile of isolation on the human proteome. Finally, we ran analyses that were adjusted for core confounders, as well as those adjusted for health behaviors. This split model adjustment was conducted as behavioral confounders may also be partial mediators of effects, with loneliness and isolation predisposing individuals to substance use, low physical activity, and other health behaviors such as sleep disturbances and poor diet (as proposed in the social control hypothesis).[18] While it is important to acknowledge their effects on molecular biomarkers, we do not know whether social deficits lead to changes in biomarkers that then causally influence health behaviors, or whether the changes in health behaviors lead to alterations in the biomarkers. Hence, we have provided results from both models.

Overall, our study showed novel protein signatures for social isolation but not loneliness in a large sample of older adults, providing the first large-scale data on the relationship between social deficits and the human proteome. Our findings demonstrated novel biological pathways influenced by social deficits including (but not limited to) those involved in cell death, malignancies, and host vulnerability, which could help to provide further biological plausibility and mechanistic evidence for the impact of social deficits on human diseases. This reinforces the importance of undertaking future research into the molecular mechanisms of social deficits.

## ONLINE METHODS

### Study population

This study used data from the English Longitudinal Study of Ageing (ELSA) - a longitudinal study of individuals in England aged 50 and older, and their partners. ELSA started by using data from the Health Survey in England (1998, 1999, and 2001) and collected its first wave of data in 2002/2003 through in-person interviews and self-reported surveys.[41] Blood samples were collected in the English Longitudinal Study of Aging (ELSA) starting from the second nurse visit in 2004 and then at four-year intervals thereafter.

The blood samples obtained during the wave 4 nurse visit in 2008 were utilized for proteomic profiling in ELSA (N = 6,271). Exclusion criteria were applied, excluding participants who either died within two years after the wave 4 nurse visit (N = 134) or were lost to follow-up (missing at least two consecutive waves) (N = 1,340). A total of 3,325 frozen plasma blood samples were sent to Olink for division into aliquots, plating, and conducting proteomics assays. A final 3,305 samples from wave 4 was viable for proteomic profiling. The analyses encompassed a final combined dataset of 3,262 participants after conducting stringent quality control of the samples (see section below). The participant selection for the proteomics assays in ELSA is depicted in Supplementary Fig. 3.

### Loneliness

Based on the wave 4 data, ELSA measured loneliness with the three-item R-UCLA loneliness scale [42, 43]: (1) how often do you feel you lack companionship? (2) how often do you feel isolated from others? (3) how often do you feel left out? Each question asks the participant to rank their experience on a three-point Likert scale (hardly ever/never, some of the time, or often), and results in a scale ranging from 3 to 9, in which a greater score corresponds with a higher level of perceived loneliness (Cronbach’s α = 0.82).

### Social Isolation

Social isolation was measured on a 7-point scale, in which each of the following represented one point: living alone, not working, not volunteering, not belonging to a club or social organisation, and having less than monthly interaction with friends, relatives, or children, respectively. This measure was adapted from the scale proposed by Bu et al.[44] The score ranged from 0 to 7, with higher value indicating greater level of social isolation.

### Proteomic data

The Olink technology employs the Proximity Extension Assay (PEA), utilizing a matched pair of antibodies labelled with unique complementary oligonucleotides (proximity probes). These probes bind to their respective target proteins in a sample, bringing them into proximity. This proximity allows the probes to hybridize, enabling DNA amplification of the protein signal. The amplified signal is then quantified using next-generation sequencing.[45]

The curation of the proteomics data in ELSA was initially to investigate proteomic signatures related to cognitive decline and dementia specifically, resulting in a more focused selection of proteins. Three Olink™ Target 96 panels were selected for the proteomics assays: Cardiovascular II (CVDII), Neurology I (NEUI), and Neurology Exploratory (NEX). These encompass an extensive array of cardiovascular, immunological and inflammatory markers, as well as markers integral to neurological processes such as axon guidance, neurogenesis, and synapse assembly.

These assays incorporate an inherent quality control mechanism utilizing four internal controls added to all samples, along with external controls. The stringent quality control pipeline has been previously described,[46] and is further detailed in Supplementary Methods.

After excluding those did not pass quality control and outliers, a final combined dataset of 3,262 samples were included in the analyses, with a total of 276 unique proteins. All proteins were quantifiable, although a total of eight proteins (BNP, MAPT, CADM3, beta-NGF, HSP90B1, NXPH1, IKZF2, EPHA10) had ≥ 50% below Limit of detection (LOD). However, these data points were not removed in the analyses, as some of the most distinct biomarkers may be low in some groups analysed but high in other groups, and by including data under LOD does commonly not increase false positives as there is generally no significant difference between groups under LOD. Protein concentrations were quantified using Olink’s standardized Normalized Protein eXpression (NPX) values, presented on a Log_2_ scale.

### Covariates

Covariates included sociodemographic factors including age, non-pension wealth quintiles, age when left formal education, sex (female vs male), and ethnicity (White vs Other ethnic group). We also included health behaviors: smoking status, alcohol consumption, physical activity, and body mass index (BMI). Participant smoking was coded to identify if an individual had never smoked, previously smoked, or currently smoked. Alcohol consumption was ordinally coded based on the number of days the participant drank alcohol in the past week (ranged from 0 to 7). Physical activity was coded as a scale ranging from participants hardly ever or never exercising, exercising monthly, exercising weekly, and exercising more than weekly. BMI was included as a continuous variable in analyses.

### Statistical analysis

#### Data normalisation and missing data handling

Missing data were addressed by imputing data using K-nearest neighbours (KNN),[47] and multiple imputation by chained equations (MICE) methods.[48] Prior to the imputation, the protein levels underwent rank-based inverse normal transformation and were scaled to achieve a mean of 0 and a standard deviation of 1. All proteins had < 6% missing. KNN imputation was used to address missing data in the proteomics data, and the number of neighbouring points (k = 57) was based on the square root of the total sample size (N = 3,262), using the ‘impute’ package in R.[49] For imputing missing data in the exposures and covariates, MICE was utilized (using the ‘mice’ R package.[50]), with 30 imputations and 10 iterations selected, to reduce bias and increase the efficiency and reliability of the statistical estimates based on empirical evidence.[51]

#### Cross-sectional linear regression model

Following this process, linear regression model was used to estimate the association between loneliness and social isolation with each protein, by pooling the estimates from all imputed datasets. Two-sided p-values were reported and displayed using a volcano plot, accompanying the coefficient, with a cut-off of 0.05 for p-value adjusted for false discovery rate (FDR; denoted as P_FDR_) to indicate statistical significance. Two multiple-adjusted linear regression models were constructed: a minimally-adjusted model in which sociodemographic variables (age, sex, ethnicity, wealth quintiles, and education) were included as covariates, and a fully-adjusted model in which sociodemographic and health behavior variables (alcohol consumption, smoking, BMI, and physical activity) variables were included as covariates.

#### Two-sample MR

Given that the limitations often associated with cross-sectional study design being unable to establish cause-and-effect relationships with temporal ambiguity, and issue with unmeasured confounding, we conducted two-sample MR on the significant proteins associated with loneliness and social isolation, based on the proteins identified from the minimally- and fully-adjusted linear regressions to infer causality,[52] by leveraging summary statistics from GWAS based on data from the UK Biobank.[53]

We identified three GWAS that were considered closely related to our **social isolation** phenotype: 1) frequency of contact with family and friends, 2) living alone, and 3) a multi-trait GWAS (MTAG) combining the results from three separate GWAS studies on perceived loneliness, living alone, and the ability to confide into a single analysis.[53] This meta-analytical approach enhances the discovery of genetic variants for a target trait by utilizing the statistical power from the additional traits. The pertinent information outlining the GWAS used for various measurements of social isolation is detailed in Supplementary Table 3.

For GWAS of protein levels, instruments representing changes in protein abundance were selected based on pQTL mapping of proteins, identifying primary genetic associations in individuals of European ancestry from the UK Biobank (https://doi.org/10.7303/syn51364943).[54] Standardization of the effects of protein pQTL was conducted to ensure alignment with the same effect allele.

Selection of instruments for exposure (various social isolation phenotypes) was carried out by considering associations at genome-wide significance (P < 5 × 10^−8^) to minimize pleiotropic effects, while excluding SNPs with a minor allele frequency (MAF) < 5%. LD (linkage disequilibrium) clumping was performed with a window size of 10,000 kilobases [kb], at an R^2^ < 0.001. In instances where a requested SNP from the exposure GWAS was not present in the outcome GWAS (protein concentration), a proxy SNP with an LD coefficient of R^2^ > 0.8 to the requested missing target SNP were sought as a substitute, utilizing the LDLink web server. LD proxies were determined using data from the 1000 Genomes European sample. The returned information included the effect of the proxy SNP on the outcome, along with details such as the proxy SNP itself, the effect allele of the proxy SNP, and the corresponding allele for the target SNP. The effects of SNPs on both outcome and exposure were then harmonized to be relative to the same allele. Positive strand alleles were inferred by utilizing allele frequencies for palindromes instead of eliminating palindromic variants. F-statistics were employed to assess the strength of SNP-exposure associations (F > 10). Effects for each individual variant were calculated using a two-term Taylor series expansion of the Wald ratio. Following this, we employed the weighted delta inverse-variance weighted (IVW) method to perform a meta-analysis of individual SNP effects, aiming to estimate the combined effect of the Wald ratios. Sensitivity analysis involved employing various MR methods, such as MR-Egger, Weighted Median, Maximum Likelihood, and Weighted Mode methods.[55] All analyses were performed using the human genome reference build GRCh38. In cases where the genome build relied on GRCh37 assembly Hg19, a lift-over process was carried out to convert genome coordinates and annotations to GRCh38 using a specified alignment, using CrossMap 0.7.0 in Python (version 3.8.8). MR analyses were conducted using the ‘TwoSampleMR’, ‘MendelianRandomization’, and ‘LDlinkR’ R packages.[56]

### Enrichment analysis

Enrichment analysis was conducted by searching open-source databases to further characterize the identified proteins based on the minimally- and fully-adjusted linear regression models. We employed Enrichr[30] using the full set of ELSA proteins as the background gene set to glean a deeper biological understanding, using Gene Ontology (GO)[57]: GO Molecular Function, GO Biological Process, and GO Cellular Component; Kyoto Encyclopaedia of Genes and Genomes (KEGG);[58] Reactome Pathway Database (REACTOME);[59] and Genotype-Tissue Expression (GTEx).[60] Statistical significance was indicated if P_FDR_ < 0.05 for the enrichment analysis. Human Protein Atlas was also searched to further characterize the identified proteins (https://www.proteinatlas.org/).[35]

All analyses were conducted using statistical software R Studio (version 4.4.0).

## ACKNOWLEDMENT

We would like to thank the participants in ELSA for their contribution to the research. We additionally want to acknowledge the participants and investigators of the UK Biobank study. We would like to thank Olink representatives and the Newcastle laboratory for their support for the proteomics data procurement and data pre-processing.

## AUTHOR CONTRIBUTIONS

The study’s conceptualization and design were undertaken by D.F., A.S., and J.G. Proteomics data curation in ELSA was a joint effort between J.G. and A.S. Z.P. handled all data processing and analysis and wrote the initial manuscript draft. D.F., A.S., and J.G. provided oversight and guidance for statistical analyses and theoretical frameworks. All authors participated in the data interpretation and subsequent manuscript’s revision process and collectively accepted responsibility for its submission for publication.

## DATA AVAILABILITY

The ELSA data is available on the UK Data Service. The proteomics data in ELSA will be deposited on the UK Data Service upon publication.

All GWAS summary statistics are available online at:

- https://doi.org/10.7303/syn51364943;
- https://gwas.mrcieu.ac.uk/;
- https://doi.org/10.17863/CAM.23511.

## CODE AVAILABILITY

The codes used for all analyses are available on GitHub repository: https://github.com/jgong94/ELSA_proteomics_SI_L.

