## Supplementary Methods, Supplementary table 1,2,3, Supplementary figure 1,2,3 for "Protein signatures associated with loneliness and social isolation: plasma proteome analyses in the English Longitudinal Study of Ageing, with causal evidence from Mendelian randomization"

### Supplementary Material

#### Supplementary Methods

##### Proteomics data quality control pipeline

*Internal quality control*

Four internal controls are added to each sample to monitor the quality of assay performance, as well as the quality of individual samples. The quality control (QC) is performed in two steps: 1. Each sample plate is evaluated on the standard deviation of the internal controls. This should be below 0.2 Normalized Protein eXpression (NPX) value. Only data from sample plate that pass this quality control will be reported; 2. The quality of each sample is assessed by evaluating the deviation from the median value of the controls for each individual sample. Samples that deviate less than 0.3 NPX from the median pass the quality control.

*External quality control*

Interquartile-range (IQR)-median and principal component analysis (PCA) analyses are used to detect and visualise outliers using ‘OlinkAnalyze’ R package. The olink_qc_plot function generates a facet plot per Panel using ggplot and ggplot2::geom_point and stats::IQR plotting IQR vs. median for all samples. This checks if any samples tend to be classified as outliers. Five standard deviations from the mean IQR and from the mean sample median are considered as outliers and excluded.

The values are by default scaled and centred in the PCA and proteins with missing NPX values are by default removed from the corresponding assay. Imputation by the median is done for assays with missingness <10% for multi-plate projects.

Based on these analyses, we then removed: 1) samples with a principal component (PC)1 or PC2 value more than five standard deviations from the mean, and 2) samples with median concentrations (NPX values) across proteins that were more than five standard deviations from the mean median, or those with IQR (NPX) across proteins that were great than five standard deviations from the mean IQR. The removal of outliers is panel specific.

**Supplementary Tables**

**Supplementary table 1. Two-sample Mendelian randomization between social isolation and protein concentration.**

| Gene name | Exposure trait | MR method | Forward MR<br>Social isolation → protein concentration<br>No of SNP | Slope (SE) | P |
|---|---|---|---|---|---|
| TNFRSF10A | Number in household | IVW | 4 | -0.511 (0.402) | 0.204 |
| TNFRSF10A | Number in household | MR Egger | 4 | -0.922 (1.182) | 0.517 |
| TNFRSF10A | Number in household | Weighted median | 4 | -0.318 (0.466) | 0.495 |
| TNFRSF10A | Number in household | Maximum likelihood | 4 | -0.516 (0.407) | 0.205 |
| TNFRSF10A | Number in household | Weighted mode | 4 | -0.214 (0.610) | 0.749 |
| TNFRSF10A | Frequency of friend/family visits | IVW | 21 | 0.245 (0.117) | 0.037 |
| TNFRSF10A | Frequency of friend/family visits | MR Egger | 21 | 1.242 (0.603) | 0.053 |
| TNFRSF10A | Frequency of friend/family visits | Weighted median | 21 | 0.324 (0.143) | 0.023 |
| TNFRSF10A | Frequency of friend/family visits | Maximum likelihood | 21 | 0.249 (0.103) | 0.016 |
| TNFRSF10A | Frequency of friend/family visits | Weighted mode | 21 | 0.357 (0.240) | 0.152 |
| TNFRSF10A | MTAG | IVW | 15 | 0.093 (0.134) | 0.488 |
| TNFRSF10A | MTAG | MR Egger | 15 | 1.086 (0.668) | 0.128 |
| TNFRSF10A | MTAG | Weighted median | 15 | 0.071 (0.179) | 0.690 |
| TNFRSF10A | MTAG | Maximum likelihood | 15 | 0.096 (0.135) | 0.476 |
| TNFRSF10A | MTAG | Weighted mode | 15 | -0.004 (0.293) | 0.990 |
| MMP12 | Number in household | IVW | 4 | 0.113 (0.418) | 0.788 |
| MMP12 | Number in household | MR Egger | 4 | 0.646 (1.490) | 0.707 |
| MMP12 | Number in household | Weighted median | 4 | 0.223 (0.482) | 0.643 |
| MMP12 | Number in household | Maximum likelihood | 4 | 0.116 (0.390) | 0.766 |
| MMP12 | Number in household | Weighted mode | 4 | 0.324 (0.687) | 0.669 |
| MMP12 | Frequency of friend/family visits | IVW | 21 | -0.053 (0.098) | 0.586 |
| MMP12 | Frequency of friend/family visits | MR Egger | 21 | -0.087 (0.529) | 0.871 |
| MMP12 | Frequency of friend/family visits | Weighted median | 21 | -0.045 (0.142) | 0.751 |
| MMP12 | Frequency of friend/family visits | Maximum likelihood | 21 | -0.052 (0.099) | 0.601 |
| MMP12 | Frequency of friend/family visits | Weighted mode | 21 | 0.226 (0.280) | 0.428 |
| MMP12 | MTAG | IVW | 15 | -0.074 (0.132) | 0.576 |
| MMP12 | MTAG | MR Egger | 15 | -0.548 (0.655) | 0.418 |
| MMP12 | MTAG | Weighted median | 15 | -0.197 (0.170) | 0.248 |
| MMP12 | MTAG | Maximum likelihood | 15 | -0.073 (0.133) | 0.587 |
| MMP12 | MTAG | Weighted mode | 15 | -0.201 (0.240) | 0.417 |

| | | | | | |
|---|---|---|---|---|---|
| TRAIL-R2 (TNFRSF10B) | Number in household | IVW | 4 | -0.295 (0.626) | 0.638 |
| TRAIL-R2 (TNFRSF10B) | Number in household | MR Egger | 4 | -2.615 (1.477) | 0.219 |
| TRAIL-R2 (TNFRSF10B) | Number in household | Weighted median | 4 | -0.532 (0.519) | 0.306 |
| TRAIL-R2 (TNFRSF10B) | Number in household | Maximum likelihood | 4 | -0.313 (0.417) | 0.454 |
| TRAIL-R2 (TNFRSF10B) | Number in household | Weighted mode | 4 | -1.033 (1.034) | 0.392 |
| TRAIL-R2 (TNFRSF10B) | Frequency of friend/family visits | IVW | 21 | 0.264 (0.102) | 0.010 |
| TRAIL-R2 (TNFRSF10B) | Frequency of friend/family visits | MR Egger | 21 | 0.359 (0.552) | 0.523 |
| TRAIL-R2 (TNFRSF10B) | Frequency of friend/family visits | Weighted median | 21 | 0.218 (0.141) | 0.123 |
| TRAIL-R2 (TNFRSF10B) | Frequency of friend/family visits | Maximum likelihood | 21 | 0.274 (0.104) | 0.008 |
| TRAIL-R2 (TNFRSF10B) | Frequency of friend/family visits | Weighted mode | 21 | 0.208 (0.258) | 0.430 |
| TRAIL-R2 (TNFRSF10B) | MTAG | IVW | 15 | 0.046 (0.137) | 0.735 |
| TRAIL-R2 (TNFRSF10B) | MTAG | MR Egger | 15 | 1.139 (0.681) | 0.118 |
| TRAIL-R2 (TNFRSF10B) | MTAG | Weighted median | 15 | 0.004 (0.184) | 0.982 |
| TRAIL-R2 (TNFRSF10B) | MTAG | Maximum likelihood | 15 | 0.047 (0.138) | 0.731 |
| TRAIL-R2 (TNFRSF10B) | MTAG | Weighted mode | 15 | -0.090 (0.380) | 0.816 |
| SKR3 (ACVRL1) | Number in household | IVW | 4 | -0.190 (0.689) | 0.782 |
| SKR3 (ACVRL1) | Number in household | MR Egger | 4 | -3.140 (1.257) | 0.130 |
| SKR3 (ACVRL1) | Number in household | Weighted median | 4 | -0.097 (0.582) | 0.868 |
| SKR3 (ACVRL1) | Number in household | Maximum likelihood | 4 | -0.203 (0.435) | 0.641 |
| SKR3 (ACVRL1) | Number in household | Weighted mode | 4 | 0.029 (0.941) | 0.978 |
| SKR3 (ACVRL1) | Frequency of friend/family visits | IVW | 21 | 0.067 (0.106) | 0.525 |
| SKR3 (ACVRL1) | Frequency of friend/family visits | MR Egger | 21 | -0.031 (0.575) | 0.958 |
| SKR3 (ACVRL1) | Frequency of friend/family visits | Weighted median | 21 | 0.031 (0.136) | 0.819 |
| SKR3 (ACVRL1) | Frequency of friend/family visits | Maximum likelihood | 21 | 0.069 (0.107) | 0.517 |
| SKR3 (ACVRL1) | Frequency of friend/family visits | Weighted mode | 21 | 0.021 (0.215) | 0.924 |
| SKR3 (ACVRL1) | MTAG | IVW | 15 | -0.087 (0.143) | 0.541 |
| SKR3 (ACVRL1) | MTAG | MR Egger | 15 | -0.018 (0.715) | 0.980 |
| SKR3 (ACVRL1) | MTAG | Weighted median | 15 | 0.020 (0.183) | 0.913 |
| SKR3 (ACVRL1) | MTAG | Maximum likelihood | 15 | -0.085 (0.144) | 0.556 |
| SKR3 (ACVRL1) | MTAG | Weighted mode | 15 | 0.199 (0.305) | 0.524 |
| TNFRSF11A | Number in household | IVW | 4 | -0.610 (0.607) | 0.314 |
| TNFRSF11A | Number in household | MR Egger | 4 | -0.420 (2.228) | 0.868 |
| TNFRSF11A | Number in household | Weighted median | 4 | -0.068 (0.501) | 0.892 |
| TNFRSF11A | Number in household | Maximum likelihood | 4 | -0.645 (0.411) | 0.116 |
| TNFRSF11A | Number in household | Weighted mode | 4 | 0.036 (0.577) | 0.955 |
| TNFRSF11A | Frequency of friend/family visits | IVW | 21 | 0.203 (0.129) | 0.116 |

| | | | | | |
|---|---|---|---|---|---|
| TNFRSF11A | Frequency of friend/family visits | MR Egger | 21 | 0.839 (0.698) | 0.244 |
| TNFRSF11A | Frequency of friend/family visits | Weighted median | 21 | 0.130 (0.151) | 0.389 |
| TNFRSF11A | Frequency of friend/family visits | Maximum likelihood | 21 | 0.210 (0.103) | 0.042 |
| TNFRSF11A | Frequency of friend/family visits | Weighted mode | 21 | -0.036 (0.300) | 0.906 |
| TNFRSF11A | MTAG | IVW | 15 | 0.026 (0.157) | 0.868 |
| TNFRSF11A | MTAG | MR Egger | 15 | 1.288 (0.760) | 0.114 |
| TNFRSF11A | MTAG | Weighted median | 15 | 0.101 (0.179) | 0.572 |
| TNFRSF11A | MTAG | Maximum likelihood | 15 | 0.027 (0.139) | 0.845 |
| TNFRSF11A | MTAG | Weighted mode | 15 | 0.115 (0.264) | 0.671 |
| VSIG2 | Number in household | IVW | 4 | 0.436 (0.428) | 0.308 |
| VSIG2 | Number in household | MR Egger | 4 | 0.397 (1.285) | 0.787 |
| VSIG2 | Number in household | Weighted median | 4 | 0.397 (0.488) | 0.415 |
| VSIG2 | Number in household | Maximum likelihood | 4 | 0.442 (0.433) | 0.307 |
| VSIG2 | Number in household | Weighted mode | 4 | 0.297 (0.776) | 0.728 |
| VSIG2 | Frequency of friend/family visits | IVW | 21 | 0.035 (0.109) | 0.749 |
| VSIG2 | Frequency of friend/family visits | MR Egger | 21 | 0.773 (0.591) | 0.207 |
| VSIG2 | Frequency of friend/family visits | Weighted median | 21 | -0.071 (0.157) | 0.649 |
| VSIG2 | Frequency of friend/family visits | Maximum likelihood | 21 | 0.036 (0.110) | 0.743 |
| VSIG2 | Frequency of friend/family visits | Weighted mode | 21 | -0.199 (0.295) | 0.508 |
| VSIG2 | MTAG | IVW | 15 | -0.261 (0.146) | 0.075 |
| VSIG2 | MTAG | MR Egger | 15 | 0.486 (0.729) | 0.517 |
| VSIG2 | MTAG | Weighted median | 15 | -0.134 (0.194) | 0.491 |
| VSIG2 | MTAG | Maximum likelihood | 15 | -0.257 (0.148) | 0.083 |
| VSIG2 | MTAG | Weighted mode | 15 | -0.033 (0.352) | 0.927 |
| PRSS8 | Number in household | IVW | 4 | -0.353 (0.430) | 0.412 |
| PRSS8 | Number in household | MR Egger | 4 | -1.070 (1.489) | 0.547 |
| PRSS8 | Number in household | Weighted median | 4 | -0.737 (0.522) | 0.158 |
| PRSS8 | Number in household | Maximum likelihood | 4 | -0.361 (0.422) | 0.392 |
| PRSS8 | Number in household | Weighted mode | 4 | -0.807 (0.711) | 0.339 |
| PRSS8 | Frequency of friend/family visits | IVW | 21 | 0.088 (0.131) | 0.500 |
| PRSS8 | Frequency of friend/family visits | MR Egger | 21 | 0.245 (0.727) | 0.739 |
| PRSS8 | Frequency of friend/family visits | Weighted median | 21 | 0.255 (0.159) | 0.110 |
| PRSS8 | Frequency of friend/family visits | Maximum likelihood | 21 | 0.092 (0.108) | 0.393 |
| PRSS8 | Frequency of friend/family visits | Weighted mode | 21 | 0.447 (0.348) | 0.214 |
| PRSS8 | MTAG | IVW | 15 | 0.137 (0.142) | 0.337 |
| PRSS8 | MTAG | MR Egger | 15 | 0.543 (0.727) | 0.468 |

| | | | | | |
|---|---|---|---|---|---|
| PRSS8 | MTAG | Weighted median | 15 | 0.306 (0.194) | 0.114 |
| PRSS8 | MTAG | Maximum likelihood | 15 | 0.142 (0.144) | 0.324 |
| PRSS8 | MTAG | Weighted mode | 15 | 0.430 (0.337) | 0.223 |
| FGFR2 | Number in household | IVW | 4 | 0.107 (0.832) | 0.897 |
| FGFR2 | Number in household | MR Egger | 4 | -3.064 (1.940) | 0.255 |
| FGFR2 | Number in household | Weighted median | 4 | -0.203 (0.578) | 0.726 |
| FGFR2 | Number in household | Maximum likelihood | 4 | 0.118 (0.442) | 0.789 |
| FGFR2 | Number in household | Weighted mode | 4 | -0.451 (0.795) | 0.610 |
| FGFR2 | Frequency of friend/family visits | IVW | 21 | 0.102 (0.113) | 0.365 |
| FGFR2 | Frequency of friend/family visits | MR Egger | 21 | 0.189 (0.628) | 0.767 |
| FGFR2 | Frequency of friend/family visits | Weighted median | 21 | 0.048 (0.156) | 0.758 |
| FGFR2 | Frequency of friend/family visits | Maximum likelihood | 21 | 0.105 (0.109) | 0.332 |
| FGFR2 | Frequency of friend/family visits | Weighted mode | 21 | -0.004 (0.277) | 0.989 |
| FGFR2 | MTAG | IVW | 15 | 0.216 (0.152) | 0.155 |
| FGFR2 | MTAG | MR Egger | 15 | 1.390 (0.716) | 0.074 |
| FGFR2 | MTAG | Weighted median | 15 | 0.334 (0.203) | 0.099 |
| FGFR2 | MTAG | Maximum likelihood | 15 | 0.221 (0.146) | 0.130 |
| FGFR2 | MTAG | Weighted mode | 15 | 0.382 (0.424) | 0.383 |
| KIM1 (HAVCR1) | Number in household | IVW | 4 | -0.346 (0.372) | 0.352 |
| KIM1 (HAVCR1) | Number in household | MR Egger | 4 | -1.124 (1.269) | 0.469 |
| KIM1 (HAVCR1) | Number in household | Weighted median | 4 | -0.470 (0.450) | 0.296 |
| KIM1 (HAVCR1) | Number in household | Maximum likelihood | 4 | -0.353 (0.377) | 0.349 |
| KIM1 (HAVCR1) | Number in household | Weighted mode | 4 | -0.590 (0.688) | 0.454 |
| KIM1 (HAVCR1) | Frequency of friend/family visits | IVW | 21 | 0.186 (0.096) | 0.052 |
| KIM1 (HAVCR1) | Frequency of friend/family visits | MR Egger | 21 | 0.621 (0.524) | 0.250 |
| KIM1 (HAVCR1) | Frequency of friend/family visits | Weighted median | 21 | 0.197 (0.133) | 0.138 |
| KIM1 (HAVCR1) | Frequency of friend/family visits | Maximum likelihood | 21 | 0.195 (0.095) | 0.040 |
| KIM1 (HAVCR1) | Frequency of friend/family visits | Weighted mode | 21 | 0.244 (0.265) | 0.368 |
| KIM1 (HAVCR1) | MTAG | IVW | 15 | -0.064 (0.158) | 0.687 |
| KIM1 (HAVCR1) | MTAG | MR Egger | 15 | -0.430 (0.801) | 0.601 |
| KIM1 (HAVCR1) | MTAG | Weighted median | 15 | -0.080 (0.172) | 0.642 |
| KIM1 (HAVCR1) | MTAG | Maximum likelihood | 15 | -0.062 (0.128) | 0.626 |
| KIM1 (HAVCR1) | MTAG | Weighted mode | 15 | -0.058 (0.254) | 0.822 |
| REN | Number in household | IVW | 4 | 0.127 (0.426) | 0.766 |
| REN | Number in household | MR Egger | 4 | 0.285 (1.325) | 0.849 |
| REN | Number in household | Weighted median | 4 | 0.155 (0.511) | 0.762 |

| | | | | | |
|---|---|---|---|---|---|
| REN | Number in household | Maximum likelihood | 4 | 0.129 (0.430) | 0.764 |
| REN | Number in household | Weighted mode | 4 | 0.219 (0.778) | 0.797 |
| REN | Frequency of friend/family visits | IVW | 21 | 0.022 (0.110) | 0.840 |
| REN | Frequency of friend/family visits | MR Egger | 21 | 0.300 (0.606) | 0.626 |
| REN | Frequency of friend/family visits | Weighted median | 21 | -0.134 (0.154) | 0.385 |
| REN | Frequency of friend/family visits | Maximum likelihood | 21 | 0.023 (0.110) | 0.833 |
| REN | Frequency of friend/family visits | Weighted mode | 21 | -0.204 (0.262) | 0.445 |
| REN | MTAG | IVW | 15 | 0.183 (0.145) | 0.207 |
| REN | MTAG | MR Egger | 15 | 0.373 (0.725) | 0.615 |
| REN | MTAG | Weighted median | 15 | 0.228 (0.199) | 0.252 |
| REN | MTAG | Maximum likelihood | 15 | 0.190 (0.147) | 0.197 |
| REN | MTAG | Weighted mode | 15 | 0.465 (0.341) | 0.194 |
| NEFL | Number in household | IVW | 4 | -0.001 (0.393) | 0.998 |
| NEFL | Number in household | MR Egger | 4 | -0.886 (1.180) | 0.531 |
| NEFL | Number in household | Weighted median | 4 | -0.051 (0.442) | 0.908 |
| NEFL | Number in household | Maximum likelihood | 4 | -0.001 (0.394) | 0.998 |
| NEFL | Number in household | Weighted mode | 4 | -0.141 (0.616) | 0.833 |
| NEFL | Frequency of friend/family visits | IVW | 21 | -0.257 (0.105) | 0.014 |
| NEFL | Frequency of friend/family visits | MR Egger | 21 | -0.068 (0.580) | 0.907 |
| NEFL | Frequency of friend/family visits | Weighted median | 21 | -0.196 (0.147) | 0.183 |
| NEFL | Frequency of friend/family visits | Maximum likelihood | 21 | -0.248 (0.101) | 0.014 |
| NEFL | Frequency of friend/family visits | Weighted mode | 21 | -0.144 (0.284) | 0.619 |
| NEFL | MTAG | IVW | 15 | -0.028 (0.161) | 0.861 |
| NEFL | MTAG | MR Egger | 15 | -0.579 (0.821) | 0.493 |
| NEFL | MTAG | Weighted median | 15 | -0.159 (0.192) | 0.406 |
| NEFL | MTAG | Maximum likelihood | 15 | -0.028 (0.136) | 0.837 |
| NEFL | MTAG | Weighted mode | 15 | -0.247 (0.270) | 0.376 |
| HAOX1 | Number in household | IVW | 4 | -0.429 (0.472) | 0.364 |
| HAOX1 | Number in household | MR Egger | 4 | -2.625 (1.301) | 0.181 |
| HAOX1 | Number in household | Weighted median | 4 | -0.170 (0.526) | 0.746 |
| HAOX1 | Number in household | Maximum likelihood | 4 | -0.441 (0.440) | 0.316 |
| HAOX1 | Number in household | Weighted mode | 4 | -0.102 (0.741) | 0.899 |
| HAOX1 | Frequency of friend/family visits | IVW | 21 | -0.086 (0.118) | 0.468 |
| HAOX1 | Frequency of friend/family visits | MR Egger | 21 | -0.047 (0.656) | 0.944 |
| HAOX1 | Frequency of friend/family visits | Weighted median | 21 | 0.047 (0.154) | 0.759 |
| HAOX1 | Frequency of friend/family visits | Maximum likelihood | 21 | -0.083 (0.111) | 0.458 |

| | | | | | |
|---|---|---|---|---|---|
| HAOX1 | Frequency of friend/family visits | Weighted mode | 21 | 0.091 (0.270) | 0.740 |
| HAOX1 | MTAG | IVW | 15 | 0.124 (0.147) | 0.399 |
| HAOX1 | MTAG | MR Egger | 15 | 0.705 (0.735) | 0.355 |
| HAOX1 | MTAG | Weighted median | 15 | 0.139 (0.206) | 0.500 |
| HAOX1 | MTAG | Maximum likelihood | 15 | 0.128 (0.149) | 0.389 |
| HAOX1 | MTAG | Weighted mode | 15 | 0.422 (0.366) | 0.268 |

*MTAG, multi-trait analysis of GWAS; MR, Mendelian randomization; SNP, single nucleotide polymorphism; SE, standard error.*

**Supplementary Table 2. Enrichment analysis for the identified proteins.**

| Term | P value | FDR-adjusted P | Genes | dataset |
|---|---|---|---|---|
| Death Receptor Activity (GO:0005035) | 0.0001 | 0.0023 | TNFRSF10B;TNFRSF10A;TNFRSF11A | GO_MF |
| CASP8 Activity Is Inhibited R-HSA-5218900 | 0.0017 | 0.0209 | TNFRSF10B;TNFRSF10A | REACTOME |
| Caspase Activation Via Death Receptors In Presence Of Ligand R-HSA-140534 | 0.0017 | 0.0209 | TNFRSF10B;TNFRSF10A | REACTOME |
| Caspase Activation Via Extrinsic Apoptotic Signaling Pathway R-HSA-5357769 | 0.0017 | 0.0209 | TNFRSF10B;TNFRSF10A | REACTOME |
| Dimerization Of Procaspase-8 R-HSA-69416 | 0.0017 | 0.0209 | TNFRSF10B;TNFRSF10A | REACTOME |
| RIPK1-mediated Regulated Necrosis R-HSA-5213460 | 0.0017 | 0.0209 | TNFRSF10B;TNFRSF10A | REACTOME |
| Regulation Of Necroptotic Cell Death R-HSA-5675482 | 0.0017 | 0.0209 | TNFRSF10B;TNFRSF10A | REACTOME |
| TP53 Regulates Transcription Of Death Receptors And Ligands R-HSA-6803211 | 0.0017 | 0.0209 | TNFRSF10B;TNFRSF10A | REACTOME |
| TRAIL Signaling R-HSA-75158 | 0.0017 | 0.0209 | TNFRSF10B;TNFRSF10A | REACTOME |
| Death Receptor Signaling R-HSA-73887 | 0.0051 | 0.0489 | TNFRSF10B;TNFRSF10A | REACTOME |
| Regulated Necrosis R-HSA-5218859 | 0.0051 | 0.0489 | TNFRSF10B;TNFRSF10A | REACTOME |
| p53 signaling pathway | 0.0017 | 0.0539 | TNFRSF10B;TNFRSF10A | KEGG |
| TP53 Regulates Transcription Of Cell Death Genes R-HSA-5633008 | 0.0099 | 0.0867 | TNFRSF10B;TNFRSF10A | REACTOME |
| Kidney - Cortex Female 60-69 Up | 0.0017 | 0.0993 | REN;HAVCR1 | GTEx |
| Kidney - Cortex Male 30-39 Up | 0.0051 | 0.0993 | REN;HAVCR1 | GTEx |
| Kidney - Cortex Male 70-79 Up | 0.0051 | 0.0993 | REN;HAVCR1 | GTEx |
| Minor Salivary Gland Female 20-29 Up | 0.0099 | 0.0993 | TNFRSF11A;PRSS8 | GTEx |
| Minor Salivary Gland Male 20-29 Up | 0.0099 | 0.0993 | MMP12;TNFRSF11A | GTEx |
| Minor Salivary Gland Male 30-39 Up | 0.0099 | 0.0993 | TNFRSF11A;PRSS8 | GTEx |
| Influenza A | 0.0099 | 0.1001 | TNFRSF10B;TNFRSF10A | KEGG |
| Natural killer cell mediated cytotoxicity | 0.0162 | 0.1001 | TNFRSF10B;TNFRSF10A | KEGG |
| Necroptosis | 0.0162 | 0.1001 | TNFRSF10B;TNFRSF10A | KEGG |
| Salmonella infection | 0.0162 | 0.1001 | TNFRSF10B;TNFRSF10A | KEGG |

| | | | | |
|---|---|---|---|---|
| Apoptosis | 0.0236 | 0.1047 | TNFRSF10B;TNFRSF10A | KEGG |
| Pathogenic Escherichia coli infection | 0.0236 | 0.1047 | TNFRSF10B;TNFRSF10A | KEGG |
| Brain - Cortex Female 50-59 Up | 0.0435 | 0.1278 | NEFL | GTEx |
| Brain - Cortex Male 30-39 Up | 0.0435 | 0.1278 | NEFL | GTEx |
| Brain - Cortex Male 70-79 Up | 0.0435 | 0.1278 | NEFL | GTEx |
| Brain - Frontal Cortex (BA9) Male 20-29 Up | 0.0435 | 0.1278 | NEFL | GTEx |
| Brain - Frontal Cortex (BA9) Male 40-49 Up | 0.0435 | 0.1278 | NEFL | GTEx |
| Brain - Frontal Cortex (BA9) Male 50-59 Up | 0.0435 | 0.1278 | NEFL | GTEx |
| Brain - Frontal Cortex (BA9) Male 60-69 Up | 0.0435 | 0.1278 | NEFL | GTEx |
| Brain - Frontal Cortex (BA9) Male 70-79 Up | 0.0435 | 0.1278 | NEFL | GTEx |
| Brain - Spinal Cord (Cervical C-1) Female 40-49 Up | 0.0435 | 0.1278 | FGFR2 | GTEx |
| Kidney - Cortex Male 40-49 Up | 0.0435 | 0.1278 | REN | GTEx |
| Kidney - Cortex Male 50-59 Up | 0.0435 | 0.1278 | REN | GTEx |
| Activation Of NF-kappaB-inducing Kinase Activity (GO:0007250) | 0.0017 | 0.1361 | TNFRSF10B;TNFRSF10A | GO_BP |
| NIK/NF-kappaB Signaling (GO:0038061) | 0.0017 | 0.1361 | TNFRSF10B;TNFRSF10A | GO_BP |
| TRAIL-activated Apoptotic Signaling Pathway (GO:0036462) | 0.0017 | 0.1361 | TNFRSF10B;TNFRSF10A | GO_BP |
| Cellular Response To Mechanical Stimulus (GO:0071260) | 0.0051 | 0.1361 | TNFRSF10B;TNFRSF10A | GO_BP |
| Extrinsic Apoptotic Signaling Pathway (GO:0097191) | 0.0099 | 0.1361 | TNFRSF10B;TNFRSF10A | GO_BP |
| Extrinsic Apoptotic Signaling Pathway Via Death Domain Receptors (GO:0008625) | 0.0099 | 0.1361 | TNFRSF10B;TNFRSF10A | GO_BP |
| Negative Regulation Of Cell-Matrix Adhesion (GO:0001953) | 0.0099 | 0.1361 | MMP12;ACVRL1 | GO_BP |
| Supramolecular Fiber Organization (GO:0097435) | 0.0162 | 0.1361 | NEFL;PRSS8 | GO_BP |
| Positive Regulation Of Transcription By RNA Polymerase II (GO:0045944) | 0.0256 | 0.1361 | MMP12;ACVRL1;FGFR2 | GO_BP |
| Activation Of Protein Kinase Activity (GO:0032147) | 0.0323 | 0.1361 | TNFRSF10B;TNFRSF10A | GO_BP |
| Apoptotic Process (GO:0006915) | 0.0323 | 0.1361 | TNFRSF10B;TNFRSF10A | GO_BP |
| Actin Filament Organization (GO:0007015) | 0.0435 | 0.1361 | PRSS8 | GO_BP |
| Activation Of Adenylate Cyclase Activity (GO:0007190) | 0.0435 | 0.1361 | PRSS8 | GO_BP |

| | | | | |
|---|---|---|---|---|
| Anterograde Axonal Transport (GO:0008089) | 0.0435 | 0.1361 | NEFL | GO_BP |
| Bone Morphogenesis (GO:0060349) | 0.0435 | 0.1361 | FGFR2 | GO_BP |
| Cardiac Muscle Tissue Morphogenesis (GO:0055008) | 0.0435 | 0.1361 | FGFR2 | GO_BP |
| Cyclic-Nucleotide-Mediated Signaling (GO:0019935) | 0.0435 | 0.1361 | PRSS8 | GO_BP |
| Defense Response To Tumor Cell (GO:0002357) | 0.0435 | 0.1361 | TNFRSF10B | GO_BP |
| Digestive System Development (GO:0055123) | 0.0435 | 0.1361 | FGFR2 | GO_BP |
| Digestive Tract Development (GO:0048565) | 0.0435 | 0.1361 | FGFR2 | GO_BP |
| Digestive Tract Morphogenesis (GO:0048546) | 0.0435 | 0.1361 | FGFR2 | GO_BP |
| Ear Morphogenesis (GO:0042471) | 0.0435 | 0.1361 | FGFR2 | GO_BP |
| Embryonic Cranial Skeleton Morphogenesis (GO:0048701) | 0.0435 | 0.1361 | FGFR2 | GO_BP |
| Embryonic Digestive Tract Development (GO:0048566) | 0.0435 | 0.1361 | FGFR2 | GO_BP |
| Embryonic Organ Morphogenesis (GO:0048562) | 0.0435 | 0.1361 | FGFR2 | GO_BP |
| Embryonic Skeletal System Morphogenesis (GO:0048704) | 0.0435 | 0.1361 | FGFR2 | GO_BP |
| Endothelial Cell Differentiation (GO:0045446) | 0.0435 | 0.1361 | ACVRL1 | GO_BP |
| Epithelial Tube Branching Involved In Lung Morphogenesis (GO:0060441) | 0.0435 | 0.1361 | FGFR2 | GO_BP |
| Epithelial Tube Formation (GO:0072175) | 0.0435 | 0.1361 | FGFR2 | GO_BP |
| Epithelium Development (GO:0060429) | 0.0435 | 0.1361 | FGFR2 | GO_BP |
| Exocrine System Development (GO:0035272) | 0.0435 | 0.1361 | FGFR2 | GO_BP |
| Forebrain Neuron Development (GO:0021884) | 0.0435 | 0.1361 | FGFR2 | GO_BP |
| Gland Morphogenesis (GO:0022612) | 0.0435 | 0.1361 | FGFR2 | GO_BP |
| Glycoprotein Catabolic Process (GO:0006516) | 0.0435 | 0.1361 | MMP12 | GO_BP |
| Inner Ear Morphogenesis (GO:0042472) | 0.0435 | 0.1361 | FGFR2 | GO_BP |
| Intermediate Filament Bundle Assembly (GO:0045110) | 0.0435 | 0.1361 | NEFL | GO_BP |
| Intermediate Filament Organization (GO:0045109) | 0.0435 | 0.1361 | NEFL | GO_BP |
| Intrinsic Apoptotic Signaling Pathway In Response To Endoplasmic Reticulum Stress (GO:0070059) | 0.0435 | 0.1361 | TNFRSF10B | GO_BP |
| Lymphatic Endothelial Cell Differentiation (GO:0060836) | 0.0435 | 0.1361 | ACVRL1 | GO_BP |

| | | | | |
|---|---|---|---|---|
| Mesenchymal Cell Differentiation (GO:0048762) | 0.0435 | 0.1361 | FGFR2 | GO_BP |
| Mesenchyme Morphogenesis (GO:0072132) | 0.0435 | 0.1361 | ACVRL1 | GO_BP |
| Morphogenesis Of An Epithelium (GO:0002009) | 0.0435 | 0.1361 | FGFR2 | GO_BP |
| Multinuclear Osteoclast Differentiation (GO:0072674) | 0.0435 | 0.1361 | TNFRSF11A | GO_BP |
| Negative Regulation Of Keratinocyte Proliferation (GO:0010839) | 0.0435 | 0.1361 | FGFR2 | GO_BP |
| Positive Regulation Of Adenylate Cyclase Activity (GO:0045762) | 0.0435 | 0.1361 | PRSS8 | GO_BP |
| Positive Regulation Of Bone Resorption (GO:0045780) | 0.0435 | 0.1361 | TNFRSF11A | GO_BP |
| Positive Regulation Of Cardiac Muscle Cell Proliferation (GO:0060045) | 0.0435 | 0.1361 | FGFR2 | GO_BP |
| Positive Regulation Of Cardiac Muscle Tissue Growth (GO:0055023) | 0.0435 | 0.1361 | FGFR2 | GO_BP |
| Positive Regulation Of Cell Division (GO:0051781) | 0.0435 | 0.1361 | FGFR2 | GO_BP |
| Positive Regulation Of Epithelial Cell Proliferation Involved In Wound Healing (GO:0060054) | 0.0435 | 0.1361 | MMP12 | GO_BP |
| Positive Regulation Of Fever Generation (GO:0031622) | 0.0435 | 0.1361 | TNFRSF11A | GO_BP |
| Positive Regulation Of Leukocyte Activation (GO:0002696) | 0.0435 | 0.1361 | HAVCR1 | GO_BP |
| Positive Regulation Of Lipase Activity (GO:0060193) | 0.0435 | 0.1361 | FGFR2 | GO_BP |
| Positive Regulation Of Mast Cell Activation (GO:0033005) | 0.0435 | 0.1361 | HAVCR1 | GO_BP |
| Positive Regulation Of Mesenchymal Cell Proliferation (GO:0002053) | 0.0435 | 0.1361 | FGFR2 | GO_BP |
| Positive Regulation Of Osteoclast Differentiation (GO:0045672) | 0.0435 | 0.1361 | TNFRSF11A | GO_BP |
| Positive Regulation Of Phospholipase Activity (GO:0010518) | 0.0435 | 0.1361 | FGFR2 | GO_BP |
| Positive Regulation Of Sodium Ion Transport (GO:0010765) | 0.0435 | 0.1361 | PRSS8 | GO_BP |
| Regulation Of Cardiac Muscle Cell Proliferation (GO:0060043) | 0.0435 | 0.1361 | FGFR2 | GO_BP |
| Regulation Of DNA Metabolic Process (GO:0051052) | 0.0435 | 0.1361 | ACVRL1 | GO_BP |
| Regulation Of Endothelial Cell-Matrix Adhesion Via Fibronectin (GO:1904904) | 0.0435 | 0.1361 | MMP12 | GO_BP |
| Regulation Of Mesenchymal Cell Proliferation (GO:0010464) | 0.0435 | 0.1361 | FGFR2 | GO_BP |
| Regulation Of Muscle Cell Differentiation (GO:0051147) | 0.0435 | 0.1361 | FGFR2 | GO_BP |
| Regulation Of Phospholipase Activity (GO:0010517) | 0.0435 | 0.1361 | FGFR2 | GO_BP |

| | | | | |
|---|---|---|---|---|
| Regulation Of Smooth Muscle Cell Differentiation (GO:0051150) | 0.0435 | 0.1361 | FGFR2 | GO_BP |
| Regulation Of Sodium Ion Transport (GO:0002028) | 0.0435 | 0.1361 | PRSS8 | GO_BP |
| Response To Lipid (GO:0033993) | 0.0435 | 0.1361 | TNFRSF11A | GO_BP |
| Response To Peptide (GO:1901652) | 0.0435 | 0.1361 | MMP12 | GO_BP |
| Retrograde Axonal Transport (GO:0008090) | 0.0435 | 0.1361 | NEFL | GO_BP |
| Skeletal System Morphogenesis (GO:0048705) | 0.0435 | 0.1361 | FGFR2 | GO_BP |
| Tumor Necrosis Factor-Mediated Signaling Pathway (GO:0033209) | 0.0435 | 0.1361 | TNFRSF11A | GO_BP |
| Vasculature Development (GO:0001944) | 0.0435 | 0.1361 | ACVRL1 | GO_BP |
| Venous Blood Vessel Development (GO:0060841) | 0.0435 | 0.1361 | ACVRL1 | GO_BP |
| Ventricular Cardiac Muscle Tissue Development (GO:0003229) | 0.0435 | 0.1361 | FGFR2 | GO_BP |
| Ventricular Cardiac Muscle Tissue Morphogenesis (GO:0055010) | 0.0435 | 0.1361 | FGFR2 | GO_BP |
| Wound Healing, Spreading Of Epidermal Cells (GO:0035313) | 0.0435 | 0.1361 | MMP12 | GO_BP |
| cAMP-mediated Signaling (GO:0019933) | 0.0435 | 0.1361 | PRSS8 | GO_BP |
| Apoptosis R-HSA-109581 | 0.0236 | 0.1439 | TNFRSF10B;TNFRSF10A | REACTOME |
| Transcriptional Regulation By TP53 R-HSA-3700989 | 0.0236 | 0.1439 | TNFRSF10B;TNFRSF10A | REACTOME |
| Cell Surface Interactions At Vascular Wall R-HSA-202733 | 0.0323 | 0.1439 | TNFRSF10B;TNFRSF10A | REACTOME |
| Programmed Cell Death R-HSA-5357801 | 0.0323 | 0.1439 | TNFRSF10B;TNFRSF10A | REACTOME |
| Assembly And Cell Surface Presentation Of NMDA Receptors R-HSA-9609736 | 0.0435 | 0.1439 | NEFL | REACTOME |
| CREB1 Phosphorylation Thru NMDA Receptor-Mediated Activation Of RAS Signaling R-HSA-442742 | 0.0435 | 0.1439 | NEFL | REACTOME |
| Cardiac Conduction R-HSA-5576891 | 0.0435 | 0.1439 | MMP12 | REACTOME |
| Degradation Of Cysteine And Homocysteine R-HSA-1614558 | 0.0435 | 0.1439 | VSIG2 | REACTOME |
| FGFR2b Ligand Binding And Activation R-HSA-190377 | 0.0435 | 0.1439 | FGFR2 | REACTOME |
| Metabolism Of Ingested SeMet, Sec, MeSec Into H2Se R-HSA-2408508 | 0.0435 | 0.1439 | VSIG2 | REACTOME |
| Muscle Contraction R-HSA-397014 | 0.0435 | 0.1439 | MMP12 | REACTOME |
| Negative Regulation Of NMDA Receptor-Mediated Neuronal Transmission R-HSA-9617324 | 0.0435 | 0.1439 | NEFL | REACTOME |

| | | | | |
|---|---|---|---|---|
| Physiological Factors R-HSA-5578768 | 0.0435 | 0.1439 | MMP12 | REACTOME |
| Ras Activation Upon Ca2+ Influx Thru NMDA Receptor R-HSA-442982 | 0.0435 | 0.1439 | NEFL | REACTOME |
| Selenoamino Acid Metabolism R-HSA-2408522 | 0.0435 | 0.1439 | VSIG2 | REACTOME |
| Signaling By FGFR2 IIIa TM R-HSA-8851708 | 0.0435 | 0.1439 | FGFR2 | REACTOME |
| Sulfur Amino Acid Metabolism R-HSA-1614635 | 0.0435 | 0.1439 | VSIG2 | REACTOME |
| Unblocking Of NMDA Receptors, Glutamate Binding And Activation R-HSA-438066 | 0.0435 | 0.1439 | NEFL | REACTOME |
| Cytokine-cytokine receptor interaction | 0.0379 | 0.1468 | ACVRL1;TNFRSF10B;TNFRSF10A;TNFRSF11A | KEGG |
| Cilium (GO:0005929) | 0.0435 | 0.1565 | HAVCR1 | GO_CC |
| Excitatory Synapse (GO:0060076) | 0.0435 | 0.1565 | FGFR2 | GO_CC |
| Intermediate Filament (GO:0005882) | 0.0435 | 0.1565 | NEFL | GO_CC |
| Intermediate Filament Cytoskeleton (GO:0045111) | 0.0435 | 0.1565 | NEFL | GO_CC |
| Motile Cilium (GO:0031514) | 0.0435 | 0.1565 | HAVCR1 | GO_CC |
| Serine-Type Peptidase Activity (GO:0008236) | 0.0236 | 0.1609 | MMP12;PRSS8 | GO_MF |
| Activin Receptor Activity (GO:0017002) | 0.0435 | 0.1609 | ACVRL1 | GO_MF |
| Adenylate Cyclase Binding (GO:0008179) | 0.0435 | 0.1609 | PRSS8 | GO_MF |
| Aspartic-Type Endopeptidase Activity (GO:0004190) | 0.0435 | 0.1609 | REN | GO_MF |
| Aspartic-Type Peptidase Activity (GO:0070001) | 0.0435 | 0.1609 | REN | GO_MF |
| BMP Receptor Activity (GO:0098821) | 0.0435 | 0.1609 | ACVRL1 | GO_MF |
| Transforming Growth Factor Beta Receptor Activity (GO:0005024) | 0.0435 | 0.1609 | ACVRL1 | GO_MF |
| Transmembrane Receptor Protein Serine/Threonine Kinase Activity (GO:0004675) | 0.0435 | 0.1609 | ACVRL1 | GO_MF |
| Tumor Necrosis Factor Receptor Activity (GO:0005031) | 0.0435 | 0.1609 | TNFRSF11A | GO_MF |

*GO_MF, Gene Ontology – Molecular Function; GTEx, Genotype-Tissue Expression; KEGG, Kyoto Encyclopedia of Genes and Genomes; REACTOME, Reactome Pathway Database. The list was filtered by P < 0.05, and terms enriched after False Discovery Rate (FDR)-correction P<0.05 were highlighted in green.*

**Supplementary Table 3. Summary information on social isolation UK Biobank GWAS datasets.**

| Trait | Number in household | Frequency of friend/family visits | MTAG of three GWAS on perceived loneliness, living alone and ability to confide |
|---|---|---|---|
| **Year** | 2018 | 2018 | 2018 |
| **Open GWAS ID** | ukb-b-5445 | ukb-b-5379 | - |
| **Consortium** | MRC-IEU | MRC-IEU | Day et al 2018 |
| **Case, n** | - | - | Loneliness: 80,134; Living alone: 2426; Ability to confide: 64,505 |
| **Control, n** | - | - | Loneliness: 364,890; Living alone: 286,524; Ability to confide: 238,062 |
| **Sample size, n** | 459,988 | 459,830 | 487,647 |
| **SNP, n** | 9,851,867 | 9,851,867 | 7,723,212 |
| **Case ascertainment** | Continuous | Categorical Ordered:<br>1 = Almost daily<br>2 = 2 – 4 times a week<br>3 = About once a week<br>4 = About once a month<br>5 = Once every few months<br>6 = Never or almost never; or no friends/family outside household | MTAG software to combine the three GWAS datasets on **perceived loneliness**, **living alone** and **ability to confide**.<br><br>**Perceived loneliness**: 'Do you often feel lonely?', as if individuals answered 'yes' (recorded as cases) or 'no' (controls).<br><br>**Living alone**: a composite variable based on the questions 'Including yourself, how many people are living together in your household?' and "How often do you visit friends or family or have them visit you?' (cases were defined as those who lived alone and who indicated that they either never visited or had no friends or family outside their household; controls were defined as those who either did not live alone or had friends who visited at least once a week).<br><br>**Ability to confide**: a variable representing the quality of social interactions 'How often are you able to confide in someone close to you?' (cases were defined as those who answered, 'Never or almost never', controls were defined as those who answered 'Almost daily'). |

*GWAS, genome wide association study; MTAG, Multi-trait analysis of GWAS; SNP, single-nucleotide polymorphism.*

#### Supplementary Figures

**Supplementary Figure 1. Volcano plot showing fully adjusted coefficient (x axis) and two-sided uncorrected P values (y axis) for the association between protein concentration with loneliness using imputed data.**

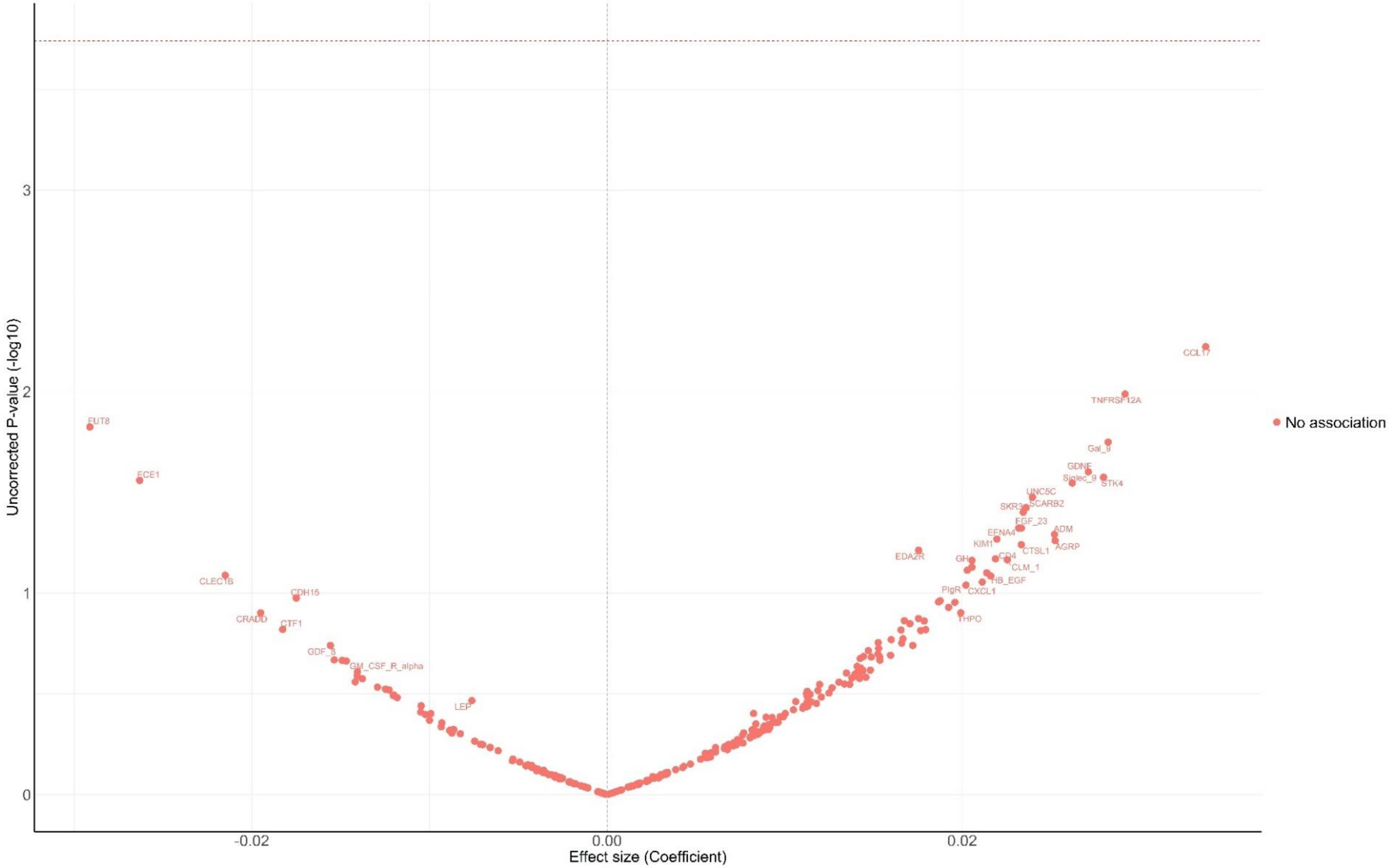


*Coefficient from linear regression models adjusted for age, sex, education, ethnicity, education, wealth quintile, alcohol consumption, smoking, body mass index, and physical activity.*

*Proteins above the horizontal dotted red line were significantly associated with loneliness FDR-corrected p-value <0.05.*

**Supplementary Figure 2. Volcano plot showing fully adjusted coefficient (x axis) and two-sided uncorrected P values (y axis) for the association between protein concentration with social isolation using imputed data.**

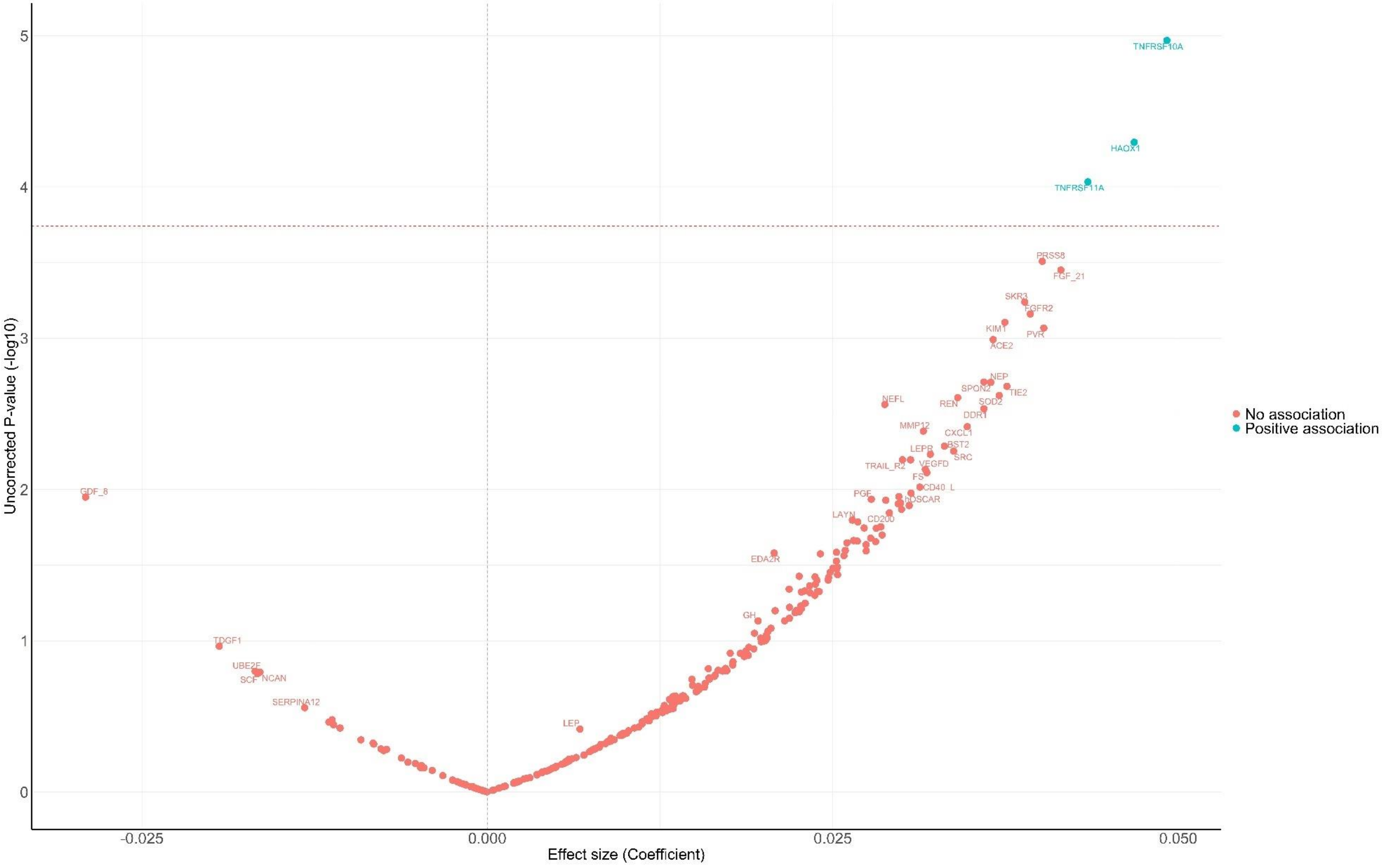


*Coefficient from linear regression models adjusted for age, sex, education, ethnicity, education, wealth quintile, alcohol consumption, smoking, body mass index, and physical activity.*
*Proteins above the horizontal dotted red line were significantly associated with social isolation FDR-corrected p-value <0.05.*

**Supplementary Figure 3. Participant selection for proteomics profiling in ELSA.**

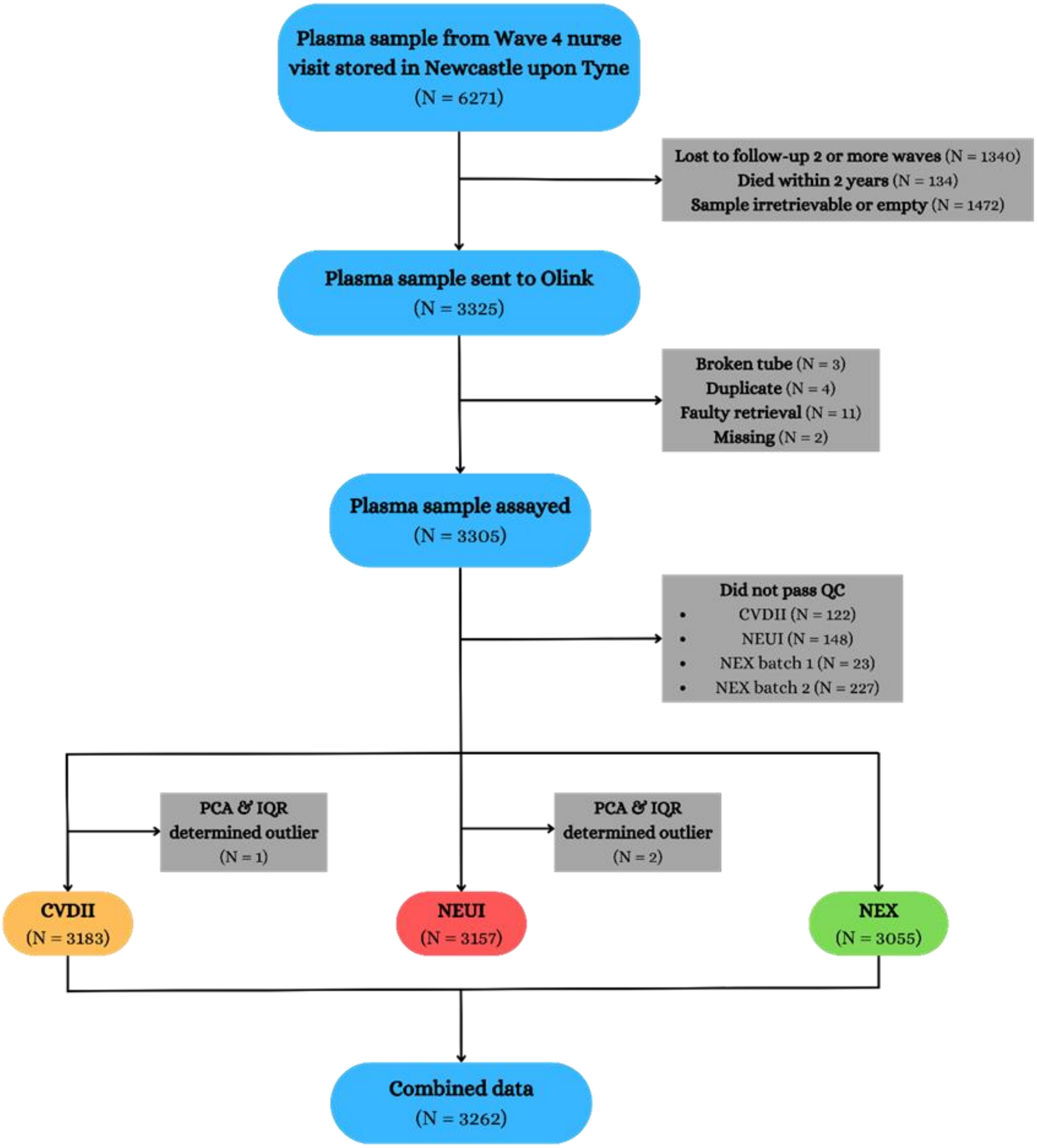


*QC, quality control; PCA, principal component analysis; IQR, interquartile range; CVDII, cardiovascular disease 2 Olink panel; NEU, neurology Olink panel; NEX, neurology exploratory Olink panel.*
